# Re-evaluating the robustness of Mendelian randomisation to measurement error

**DOI:** 10.1101/2022.10.02.22280617

**Authors:** Benjamin Woolf, James Yarmolinsky, Ville Karhunen, Kate Tilling, Dipender Gill

## Abstract

**Background:** Mendelian randomisation (MR) uses germline genetic variation as a natural experiment to investigate causal relations between traits. MR is robust to non-differential random measurement error in exposures or outcomes. However, the effect of differential measurement error, and non-differential measurement error on the variant selection process, remains unclear.

**Methods:** We use Monte-Carlo simulations and an applied example to explore the effect of differential measurement error on MR estimates for a continuous exposure and outcome, and the application of multivariable MR to reduce bias. We then explore the effect of non-differential measurement error during variant selection on MR analysis, using simulated and real-world data in the UK Biobank.

**Results:** Causal differential measurement error biased MR estimates when it occurred in the outcome, or in an exposure with a true causal effect on the outcome. This bias was mitigated by including the variable causing the error in a multivariable MR analysis. Unlike standard regression, MR was not biased by non-causal differential measurement error, i.e. when a third variable caused the exposure (or outcome) and the error in the outcome (or exposure). Non-differential measurement error in the phenotype during variant selection reduced the precision of MR estimates and induced bias. This bias was attenuated by using three-sample MR, or Winner’s curse corrections.

**Conclusion:** MR estimates can be biased by differential measurement error, but in fewer circumstances than standard regression. Multivariable MR can be used to attenuate differential measurement error if the error mechanism is known. Three-sample MR is recommended particularly for error-prone exposures.

**Key Messages:**

- Previous research demonstrates that Mendelian randomization (MR) is unbiased by (classical) non-differential measurement error in the exposure or outcome once the genetic instruments have been identified.
- MR estimates can be biased by causal differential measurement error in a continuous outcome, or in a continuous exposure when there is a true causal effect of the exposure on the outcome. As with observational studies, this bias could lead to an over-or under-estimation of the true effect estimate.
- Unlike standard regression, MR is not biased by non-causal differential measurement error between the exposure and outcome, or causal differential measurement error in the exposure under the null hypothesis.
- When all the requisite assumptions are met, multivariable MR can be used to attenuate bias due to differential measurement error in an exposure or outcome, if the variables causing the error are known. Else, a smaller sample, which is less susceptible to differential measurement error, would produce more accurate estimates, despite decreased power.
- Non-differential measurement error in the exposure will reduce precision and can cause bias in MR when it occurs during the instrument selection process. The bias caused by non-differential measurement error in instrument selection can be mitigated by using non-overlapping samples for instrument selection and the instrument-exposure estimation, or statistical correction for Winner’s curse.

## Introduction

Mendelian randomisation (MR) is a method used to estimate the causal effect of a modifiable exposure on an outcome, under specific assumptions.(1–3) In MR, confounding bias is avoided by using genetic variants (usually single-nucleotide polymorphisms [SNPs]) associated with an exposure of interest as instrumental variables (IV) for the exposure.(4) The causal effect can be estimated in one sample (one sample MR, or 1SMR), or separately estimated using two separate samples (two-sample MR, or 2SMR).(3,5) Variants to be used as instruments may be selected based on a statistical criterion, such as a genome-wide significant p-value from a genome-wide association study (the “discovery” GWAS).. It is advisable to use a separate GWAS to estimate the associations between variants and the exposure (three sample MR, or 3SMR), to avoid bias from the “Winner’s curse”.(6) However, this is not common in practice.(7)

A systematic review of the reporting of two-sample MR studies found that only 1% of the papers examined the possible impact of measurement error.(7) Measurement error occurs when there is a difference between the true and measured values, and can be described as either differential (error differs according to some participant characteristics) or non-differential.(8) Classical measurement error is when the observed variable is an unbiased proxy for the true variable. Differential measurement error will bias effect estimates in a linear regression model when the error in the exposure correlates with the outcome, or when the error in the outcome correlates with the exposure.(9,10) Because of this, differential measurement error is often defined in terms of the analysis model.(11) In a linear regression model, classical non-differential (random) measurement error will reduce precision when it occurs in the outcome, and bias estimates towards the null when it occurs in the exposure.(12,13).

Simulations of MR, and instrumental variable (IV) analyses more generally, have shown that (classical) non-differential measurement error in the exposure or outcome has a minimal effect on the precision of traditional IV estimators, and does not bias estimates towards the null.(14–17) This may explain the low proportion of papers discussing measurement error as a potential threat to validity, and has resulted in MR being described as being robust to measurement error.(18,19) However, the extent to which MR is affected by differential measurement error is unclear.

We are aware of no studies that have explored the impact of non-differential measurement error in the discovery GWAS. Non-differential measurement error in the discovery sample could affect MR in two ways. First, the power of the GWAS, and therefore the number of true SNPs detected, will be decreased, which will reduce the power of the MR study.(20) Second, the reduced power of the discovery GWAS will increase the chance that a SNP detected will be a false positive, therefore increasing the risk of Winner’s curse.(6,21)

This paper aims to explore the impact on MR analyses of differential measurement error in the exposure or outcome on MR analyses, and phenotypic non-differential measurement error in the GWAS discovery study.

## Methods

### Overview of the study design

Firstly, we use a Monte-Carlo simulation to explore the effect of differential measurement error in the exposure and outcome on one and two-sample MR estimates. We then use Monte-Carlo simulations to examine the conditions under which multivariable MR (MVMR) can be used to attenuate bias when the variable causing the measurement error is known and has a valid instrument. Next, we provide an applied example looking at the effect of body mass index (BMI) on fasting and non-fasting blood glucose. Finally, we use Monte-Carlo simulation and an applied example of the effect of bodyweight on cardiovascular disease (CVD) risk, to examine the impact of non-differential measurement error in the discovery GWAS on MR estimates. We examine the use of a Winner’s curse correction, or estimating the SNP-exposure associations in external data, as methods to attenuate bias.

### Measurement error mechanisms considered

We explore four types of differential measurement error (Scenarios A-D in Table 1). Outcome-related measurement error in the exposure is when the participants’ outcome status associates with measurement error in the exposure. Exposure-related measurement error in the outcome is defined as when the participants’ exposure status associates with measurement error in the outcome. Each of these error mechanisms could be direct (exposure directly causes measurement error in the outcome), indirect (exposure causes variable N which causes measurement error in the outcome), or confounded (variable U causes exposure and also causes measurement error in the outcome).

**Table 1:**
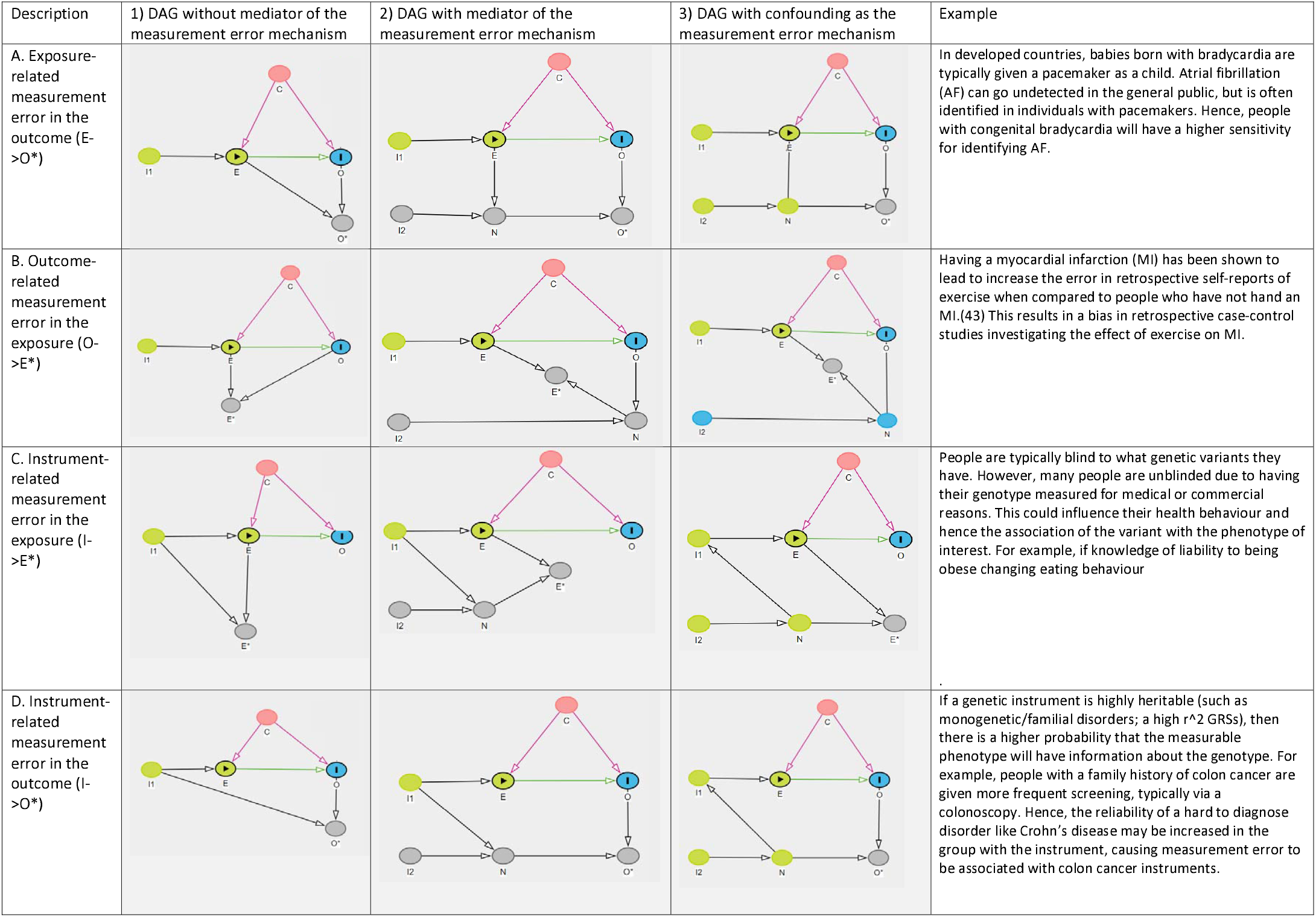
DAGs of causal structures used to simulate differential measurement error. In each DAG ‘I1’ and ‘I2’ represents the genetic instruments, ‘N’ is a noise variable which mediates the biasing pathway, ‘E’ the exposure, ‘C’ is a confounder, and ‘O’ is the outcome. ‘E*’ and ‘O*’ are the observed (measured with error) exposure and outcomes respectively. DAG: directed acyclic graph.

| Description | 1) DAG without mediator of the measurement error mechanism | 2) DAG with mediator of the measurement error mechanism | 3) DAG with confounding as the measurement error mechanism | Example |
| --- | --- | --- | --- | --- |
| A. Exposure-related measurement error in the outcome ($E \rightarrow O^*$) | | | | In developed countries, babies born with bradycardia are typically given a pacemaker as a child. Atrial fibrillation (AF) can go undetected in the general public, but is often identified in individuals with pacemakers. Hence, people with congenital bradycardia will have a higher sensitivity for identifying AF. |
| B. Outcome-related measurement error in the exposure ($O \rightarrow E^*$) | | | | Having a myocardial infarction (MI) has been shown to lead to increase the error in retrospective self-reports of exercise when compared to people who have not had an MI.(43) This results in a bias in retrospective case-control studies investigating the effect of exercise on MI. |
| C. Instrument-related measurement error in the exposure ($I \rightarrow E^*$) | | | | People are typically blind to what genetic variants they have. However, many people are unblinded due to having their genotype measured for medical or commercial reasons. This could influence their health behaviour and hence the association of the variant with the phenotype of interest. For example, if knowledge of liability to being obese changes eating behaviour |
| D. Instrument-related measurement error in the outcome ($I \rightarrow O^*$) | | | | If a genetic instrument is highly heritable (such as monogenetic/familial disorders; a high $r^2$ GRSs), then there is a higher probability that the measurable phenotype will have information about the genotype. For example, people with a family history of colon cancer are given more frequent screening, typically via a colonoscopy. Hence, the reliability of a hard to diagnose disorder like Crohn's disease may be increased in the group with the instrument, causing measurement error to be associated with colon cancer instruments. |

Instrument-related measurement error in the outcome is defined as when the genotype influences measurement error in the outcome, and instrument-related measurement error in the exposure is defined as when the genotype influences the measurement error in the exposure. For example, a previous MR study used an increased risk of gallbladder cancer among genetically predicted gallstones cases to argue for a causal effect of having gallstones on gallbladder cancer.(22) However, most of the gallbladder cancer cases diagnoses were made after initiating prophylactic therapy to prevent gallstones. One alternative explanation is that the increased surveillance in people with a higher genetic risk for gallstones resulted in more accurate detection of gallbladder cancer in gallstone cases than controls.

### Impact of differential measurement error in the exposure or outcome on estimates of the causal effect of exposure on outcome from MR or linear regression

We used the Directed Acyclic Graphs (DAGs) in Table 1 (scenarios A-D) to explore the impact of each mechanism on likely bias in the causal estimate. We then used these DAGs (Table 1, scenarios A1-D1) as the framework to simulate data on instrument, exposure, outcome and observed (measured with error) exposure and outcome. We conducted our simulations using the ADEMP (aims, data-generating mechanisms, estimates, methods, and performance measures) approach, with details in Section 1 of the Supplementary Methods.(23)

### Use of multivariable MR (MVMR) to reduce bias due to differential measurement error in the exposure or outcome

MVMR is an extension of multivariable IV methods to MR.(24,25) An MVMR analysis in which the IV model includes more than one exposure and instrument will estimate the direct effect of each exposure on the outcome adjusted for the other exposures.

Where researchers understand the path through which the differential measurement error occurs and have measured a variable that mediates the measurement error mechanism, it should be possible to attenuate the differential error by including both the exposure and the measurement error mechanism in an MVMR model. We used DAGs to examine the theoretical conditions under which the bias would be removed when the measurement error was entirely mediated by a known variable with a valid instrument, in Scenarios A2-D2 (Table 1). We then used these DAGs as the basis of a simulation to explore the use of MVMR to attenuate bias due to differential measurement error. We additionally explored the robustness of this method to violations of the MVMR assumptions, including via an error-prone measure of the mediating variable, and population stratification, and when the measurement error mechanism occurred through confounding rather than mediation. Further information on these analyses is presented in Section 2 of the Supplementary Methods.

### Applied example of differential measurement error in the effect of BMI on fasting and non-fasting blood glucose

Individual characteristics can influence the measurement of blood metabolites, such as blood glucose. It is therefore typical for studies to require that participants have fasted for a pre-set period before blood samples are collected.(29,30) However, the UK Biobank (UKB) measured non-fasting serum levels. This may make these measures more susceptible to differential measurement error.

Higher BMI may impact baseline blood glucose levels directly or through diseases like diabetes. However, BMI also has implications for appetite and food consumption, and when we eat our measured blood glucose level (i.e. non-fasting blood glucose) typically increases. Thus measuring the effect of BMI on baseline blood glucose using non-fasting samples may introduce a differential measurement error. Because fasting blood sugar is generally higher than non-fasting blood sugar levels, there will likely also be a difference in the mean value of the two measures (Figure 1).

**Figure 1:**
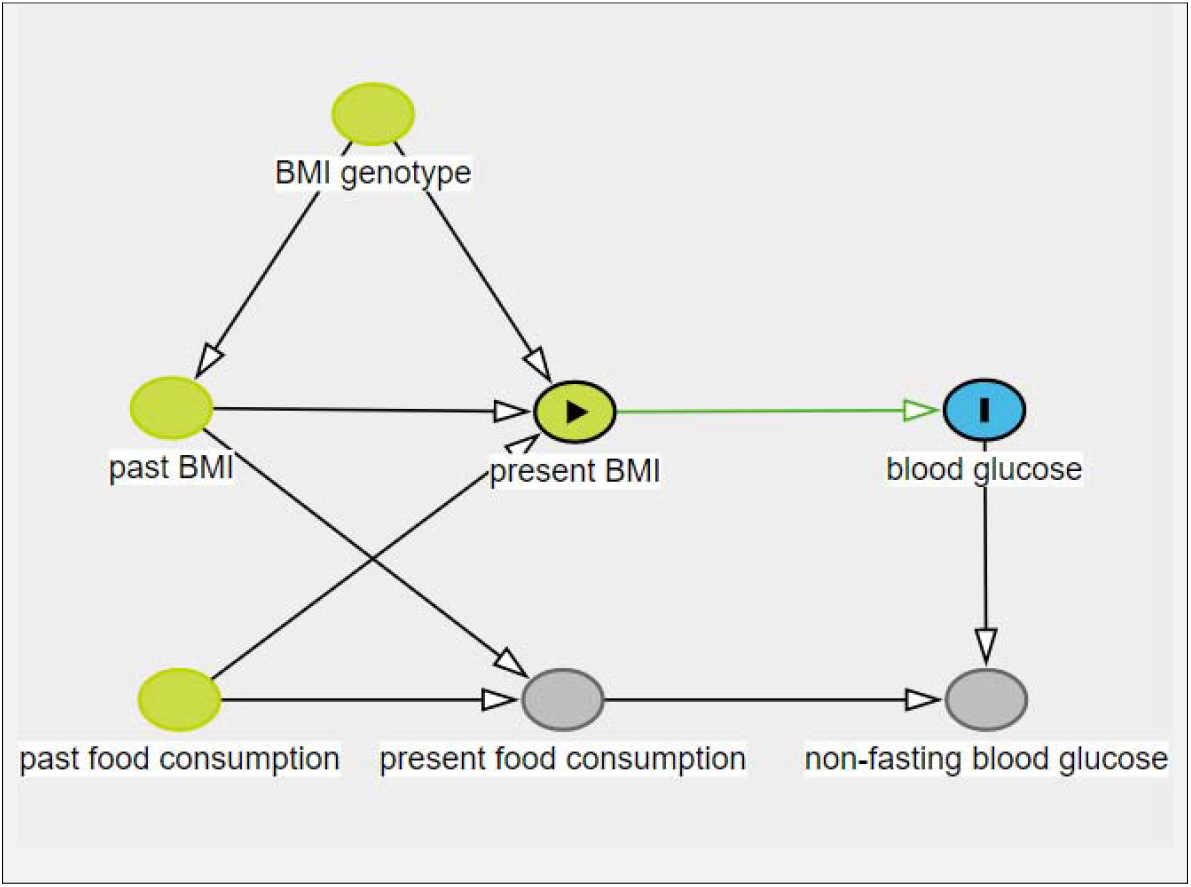
DAG representation of how (non-fasting) blood glucose levels can be differentially biased by BMI. In this DAG, present BMI is the exposure and (fasting/baseline) blood glucose is the outcome. Blood glucose is measured through a proxy, non-fasting blood glucose. However, because having a higher BMI increases one’s food consumption in the present, which in turn increases non-fasting blood glucose, differential measurement error could introduce. DAG: directed acyclic graph; BMI: body mass index.

We attempted to quantify any such bias using a two-sample MR design. In brief, we used the GIANT consortium BMI GWAS (339,224 participants, 95% of European ancestry) to estimate the SNP-exposure associations, with the MAGIC fasting blood glucose (133,010 European ancestry participants),(10) and the UKB non-fasting blood glucose (114,867 predominantly of European ancestry participants) GWAS to estimate the SNP-outcome associations.(32–36) The UKB did not overlap with either MAGIC or GIANT, although the latter two shared a maximum of 370,000 participants (i.e. 28% and 11% respectively, Supplementary Table 1). Neither blood glucose GWAS adjusted for BMI.

On the same visit as the non-fasting blood glucose samples were measured, the UKB asked participants how long it had been since they last consumed any food or drink, and how much food they had eaten the previous day. We used UKBB to identify instruments for time since they had last consumed food or drink, and for food weight eaten on the previous day, and used them. We then explored the use of (one-sample) MVMR in attenuating bias by adjusting for time since food or drink consumption.

Further details on these analyses can be found in Section 6 to 11 in the Supplementary Methods.

### Impact of phenotypic non-differential measurement error in the discovery sample on MR estimates

#### Simulation study

We simulated a series of discovery GWASs with increasing amounts of measurement error in the phenotype, and then used them to select instruments for a simulated MR. The primary estimands were the bias of causal effect, and the standard error (SE) of the estimated effect, of the exposure on the outcome when:

1. The same sample was used to estimate the variant-exposure association as to select variants.
2. The same sample was used but the False discovery rate Inverse Quantile Transformation (FIQT) Winner’s curse correction was applied as a method of addressing any potential bias. This correction uses an analogy between Winner’s curse and multiple testing as the basis for an easy to implement correction (see Section 5 of the Supplementary Methods for more details).
3. Variant selection and effect estimation occurred in separate samples. More details can be found in Section 3 of the Supplementary Methods.

#### UKB Analysis

We used a two-sample MR design, with and without the FIQT correction to examine the effect of having a higher bodyweight on CVD risk. For a discovery sample, we used GWAS of bodyweight in the UK Biobank (each with 461,632 mostly European ancestry participants). We ran a total of 106 GWAS. Non-differential measurement error was increased by adding a random normal variable with a mean of zero and incrementally larger standard deviations. As a source of CVD risk genetic associations, we used the FinnGen CVD GWAS (111,108 cases and 107,684 controls from the Finnish population). A GIANT GWAS of bodyweight (133,723 European participants) was used as a replication GWAS for the exposure for the three sample MR setting. Further details can be found in Sections 6 to 8 of the Supplementary Methods.

## Results

### Impact of differential measurement error in the exposure or outcome on MR and linear regression estimates of the causal effect

From the DAG for scenario A1 (Table 1), the systematic error in the exposure that is caused by the outcome induces a backdoor path from the observed exposure to outcome. Thus, a linear regression of outcome on observed exposure would give a biased estimate of the causal effect of exposure on outcome. Similarly, there is a path from instrument to true exposure to outcome and then to observed exposure, which means that the instrument-exposure estimate will be biased, and this will bias the MR estimate of the causal effect. However, the instrument-outcome association is not biased, and thus if exposure does not cause outcome, the estimated causal effect using MR will not be biased.

From the DAG for scenario B1 (Table 1), systematic error in the outcome that is caused by the exposure induces a spurious direct path from exposure to observed outcome. Thus, a linear regression of observed outcome on exposure would give a biased estimate of the causal effect of exposure on outcome. Similarly, there is a path from instrument to exposure to observed outcome, which means that the MR estimate of the causal effect will be biased. The path causing the bias does not include the exposure-outcome effect, and thus the bias will occur even under the null. The DAGs for an indirect mechanism via variable N (scenarios A2 and B2, Table 1) lead to the same bias conclusions as above.

However, the DAGs for a non-causal mechanism via U which causes exposure/outcome and error in outcome/exposure (scenarios A3 and B3, Table 1) lead to differing conclusions about bias in MR or linear regression. For MR, these two scenarios do not lead to bias, irrespective of whether there is in truth a causal effect of exposure on outcome. For linear regression analyses, they both lead to bias. This is because this non-causal mechanism essentially introduces confounding between the observed exposure and outcome, which does not lead to bias when using MR.

From the DAGs for instrument-related error in exposure or outcome (scenarios C1 and D1, Table 1), the error mechanism induces a spurious path from the instrument to observed exposure (C1) or from instrument to outcome (D1). Thus, the MR analyses will be biased in each case. If there is in truth no causal effect of exposure on outcome, the instrument-related error in the exposure will not bias the MR estimate, as the instrument to outcome pathway is not biased. The instrument-related error in the exposure (C1) causes bias in the linear regression estimate of the causal effect as it induces a backdoor bath from observed exposure to outcome via the true exposure. As with MR, this bias will not occur under the null. The instrument-related error in the outcome (D1) also causes bias in the linear regression estimate of the causal effect, as it effectively adds confounding by the instrument of the exposure-outcome association.

Our simulations confirmed the conclusions from the DAGs above (Supplementary Results Section 1, and Supplementary Table 3).

### Examining the use of multivariable MR to reduce bias due to differential measurement error in the exposure or outcome

The DAGs for all scenarios other than A2 show that conditioning on N (whether via multivariable linear regression or via MVMR) will block the error mechanism and therefore remove the bias. In scenario A2, the bias towards the null that results from the variance in the observed exposure not being the same as that for the true exposure is not removed by conditioning on N, and therefore the multivariable linear regression estimate of the direct effect of exposure on outcome is still biased. The MVMR estimate is not biased, in the same way that MR is not biased by random measurement error in the exposure.

On a practical note, if there are confounders of N and the outcome, then multivariable linear regression (conditioning on N) will not be unbiased, due to the well-known effect of conditioning on a mediator in the presence of mediator-outcome confounding.

### Simulations exploring the use of multivariable MR to reduce bias due to differential measurement error in the exposure or outcome

Our simulation found that when the variable inducing the measurement error mechanism had been measured, and has a valid instrument, MVMR gives an unbiased estimate of the true causal effect, with nominal coverage (Supplementary Table 4). MVMR generally failed to attenuate bias and had reduced coverage when the MVMR assumptions were violated, or when the measured variable only partially mediated the differential measurement error (Supplementary Table 5). MVMR did attenuate bias and restore the 95% confidence intervals in the simulations where classical measurement error was added to the measurement error mechanism, and when the measurement error mechanism occurred through confounding rather than mediation.

### Applied example of differential measurement error comparing the effect of BMI on fasting/non-fasting blood glucose

Using non-fasting rather than fasting blood glucose as an outcome in two-sample MR resulted in estimates that were approximately 25% further from the null. For example, the IVW estimate for the effect of a standard deviation increase of BMI on non-fasting and fasting blood glucose was 0.103 mmol/L (95% CI: 0.058 to 0.148) and 0.074 mmol/L (95% CI: 0.045 to 0.103)) respectively (Figure 2). In the 1SMR setting, we found a similar (24%) change from 0.179 (19% CI 0.152 to 0.206) mmol/L to 0.136 (95% CI: 0.065 to 0.207) mmol/L for the effect of BMI on non-fasting glucose in a univariable analysis compared to the MVMR estimate after adjusting for the two food consumption variables. Supplementary Table 2 presents a set of potential alternative explanations for why the one-sample and two-sample MR estimates differ.

**Figure 2:**
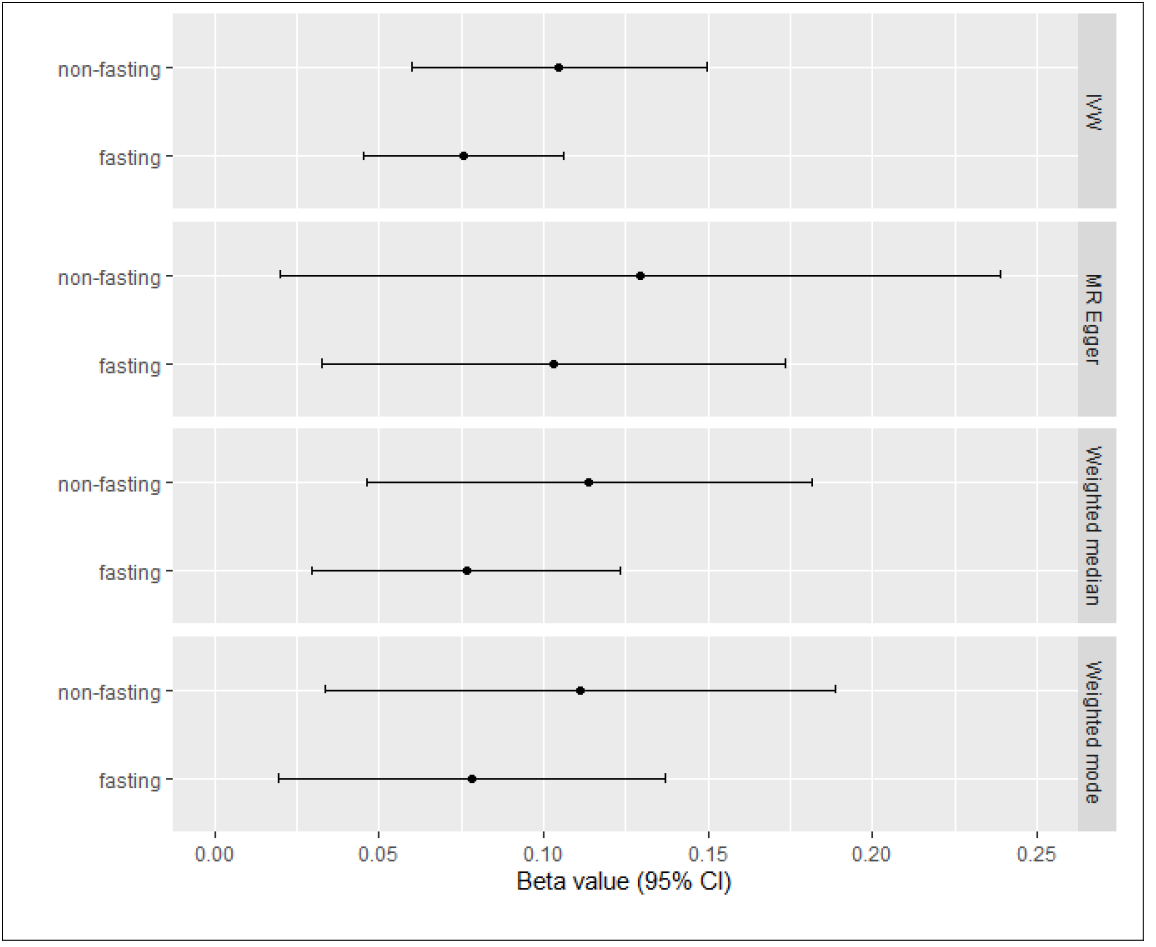
Mendelian randomisation (MR) effect size estimates of genetically proxied body mass index (BMI) on fasting and non-fasting blood glucose. IVW: inverse-variance weighted method. CI: confidence interval.

### Phenotypic non-differential measurement error in the discovery GWAS

Increasing phenotypic measurement error in the discovery GWAS reduced the number of selected SNPs (Supplementary Figure 2), and increased the standard error of the MR estimates (Supplementary Figure 3), in both the real and simulated data settings. Increasing measurement error biased both the variant-exposure associations, and MR estimates, when variant selection and variant-exposure effect estimation occurred in the same sample (Supplementary Figure 4, Figure 3). The FIQT Winner’s curse correction attenuated the bias; and it was mitigated completely by using three samples. Further details can be found in the Supplementary Results.

**Figure 3:**
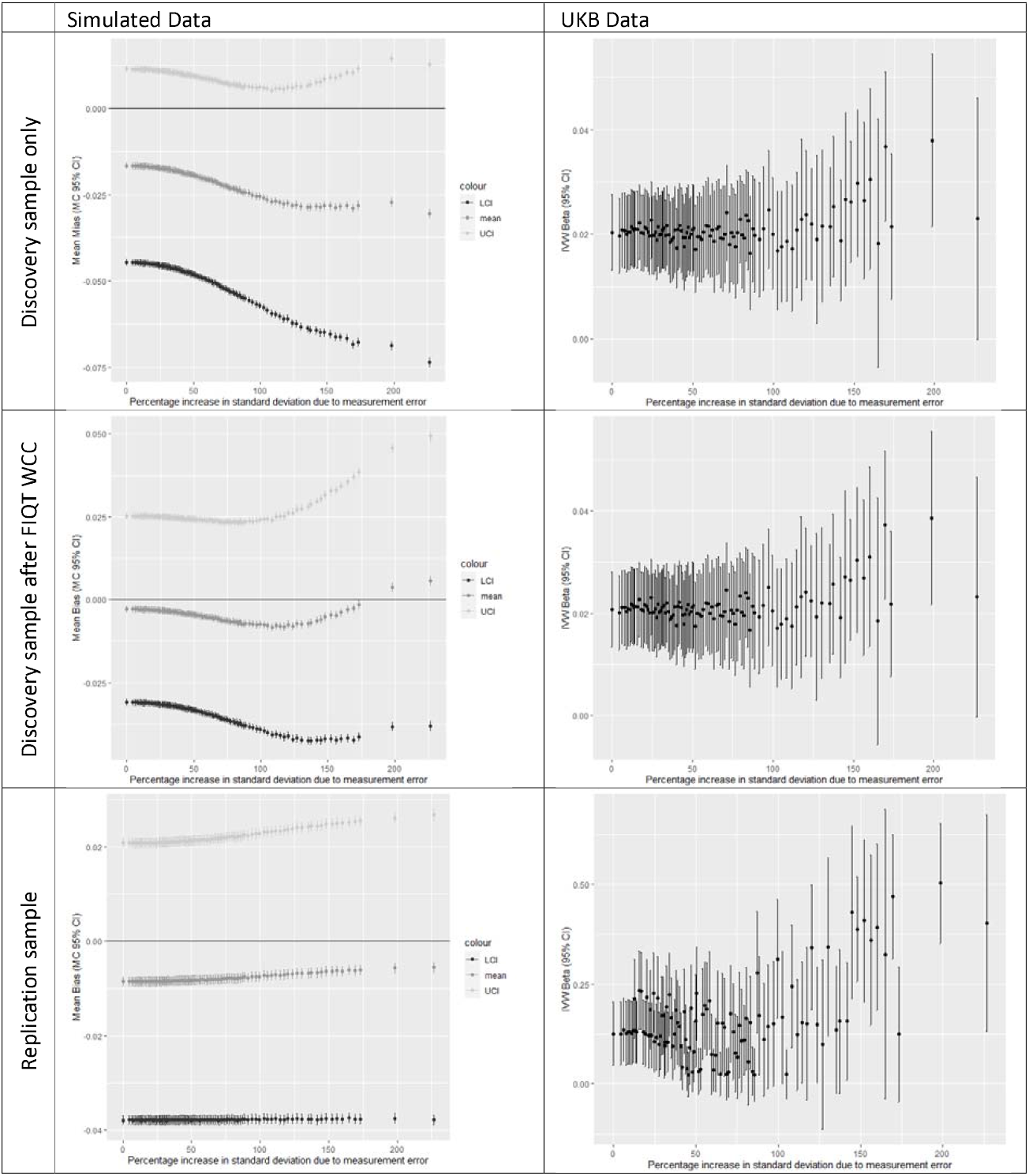
The impact of non-differential measurement error in the discovery sample on the inverse-variance weighted (IVW) Mendelian randomisation estimates. MC: Monte Carlo; CI: confidence interval; IQR: interquartile range; FIQT WCC: False discovery rate Inverse Quantile Transformation Winners Curse Correction.

## Discussion

### Key Results and Interpretation

MR is not biased by (classical) non-differential measurement error in the exposure or outcome, providing that strong and valid instruments are used.(14–17) Here we show that causal differential error in the outcome or exposure may bias MR estimates. However, differential measurement error in the exposure did not cause bias when there truly was no effect of the exposure on outcome. When the measurement error mechanism was non-causal (e.g., it occurs through a confounder), differential measurement error in the exposure (or outcome) due to the outcome (or exposure) did not bias MR. Standard regression analyses would be biased by both causal and non-causal differential measurement error mechanisms, even under the null. If the variables causing error in the exposure or outcome are known and can be instrumented, then MVMR can be used to attenuate bias due to this differential measurement error. However, the estimates from MVMR may address a different causal question, because the estimate is conditional on the other phenotypes adjusted for.

Non-differential measurement error biases estimates in the discovery sample (Figure 3), but not in the replication sample. This demonstrates the importance of using “three-sample MR”, with separate samples for choosing which SNPs to use as instruments and for estimating the SNP-exposure association (as others have already argued (6,38)). When this is not possible, we have shown that Winner’s curse corrections, like the FIQT, can be used to attenuate this measurement error bias. The comparatively small amount of bias caused by simulated phenotypic measurement error in the discovery sample of UKB implies that Winner’s curse from measurement error can also be reduced by using very large samples. However, it will be difficult to know in practice if the sample size is sufficiently large to counteract any bias from non-differential measurement error. One implication of these findings is that selecting GWASs for use in an MR analysis based on their sample size alone may be a sub-optimal strategy. Our example using fasting glucose showed that researchers may have to trade off the potential merits of a larger GWAS with that of using data from a smaller sample with less error-prone measures.

### Strengths and Limitations

We only explored two measurement error mechanisms (classical measurement error in a normally distributed variable, and differential error due to the linear effect of another variable).(15) Other types of measurement error not modelled here, including non-linear/multiplicative errors, parallax, or calibration error, may behave differently to the types of error we have considered.(17) We have only examined continuous exposure and outcome, and have not examined misclassification of binary or categorical exposures or outcomes. We also did not explore all the possible ways in which measurement error could arise in the discovery, replication, or outcome data set in MR.

A limitation of the use of MVMR to adjust for differential measurement error is that it requires the ability to identify and instrument the measurement error mechanism. In our example, we did not have very strong instruments for our proposed causes of measurement error (e.g., duration of time since eating/drinking), and thus the MVMR assumptions may not have been met.

Pleiotropy robust methods are unlikely to be a viable solution for attenuating any bias due to differential measurement error because their relaxed assumptions are also likely to be violated. For example, all four types of differential measurement error explored in our simulations should exert a homogeneous bias across all SNPs. Thus, methods like MR-Mode, MR-Median and MR-PRESSO, which assume the validity of non-outlier SNPs, will still be biased.

The development of largescale publicly available omics data repositories, such as the MRC-IEU OpenGWAS project,(37) or the NHGRI-EBI GWAS Catalogue,(42) may facilitate the identification of alternative data sources. Because it is unlikely that any one source of data will have the best quality phenotyping and the best power, we would suggest that authors take the time to consider the merits of multiple data sources for each phenotype, and, if needed, attempt to triangulate across them. Finally, one potential advantage of using a two-sample MR design is that by measuring the exposure(s) and outcome in independent samples, knowledge about the measurement of the exposure may be less likely to influence the measurement or the outcome, and vice versa, than in a study in which exposure and outcome status is measured in the same participants and/or by the same study personnel.

## Supporting information

Supplementary Results

## Data Availability

STROBE-MR extension

## Additional Information

### Funding

Benjamin Woolf is funded by an Economic and Social Research Council (ESRC) South West Doctoral Training Partnership (SWDTP) 1+3 PhD Studentship Award (ES/P000630/1). James Yarmolinsky is supported by a Cancer Research UK Population Research Postdoctoral Fellowship (C68933/A28534). DG is supported by the British Heart Foundation Centre of Research Excellence at Imperial College London (RE/18/4/34215) and by a National Institute for Health Research Clinical Lectureship at St. George’s, University of London (CL-2020-16-001). BW, KT and JY work in the MRC Integrative Epidemiology Unit that is supported by the University of Bristol and UK Medical Research Council (MC_UU_00011/1, MC_UU_00011/3, MC_UU_00011/7), and the CRUK-funded Integrative Cancer Epidemiology Programme (C18281/A1916). VK is funded by the Academy of Finland Project 326291, and European Union’s Horizon 2020 research and innovation programme under Grant Agreement No 848158 (EarlyCause). DG is funded by the British Heart Foundation Centre of Research Excellence (RE/18/4/34215) at Imperial College London. This research was funded by United Kingdom Research and Innovation Medical Research Council (MC_UU_00002/7). For the purpose of open access, the author has applied a Creative Commons Attribution (CC BY) licence to any Author Accepted Manuscript version arising from this submission.

### Contributions

BW conceived of the study with the help of JY and DG. BW and DG designed the analyses on differential measurement error, KT and BW designed the analyses on non-differential measurement error in the exposure. BW conducted the analysis and drafted the manuscript. All authors provided feedback on the analysis design and manuscript and contributed to the interpretation of the results.

### Data and data sharing

This project was conducted using UK Biobank application no. 15825. UK Biobank was established by the Wellcome Trust medical charity, Medical Research Council, Department of Health, Scottish Government and the Northwest Regional Development Agency. It has also had funding from the Welsh Government, British Heart Foundation, Cancer Research UK and Diabetes UK. UK Biobank is supported by the National Health Service (NHS). UK Biobank is open to bona fide researchers anywhere in the world.

This work was carried out using the computational facilities of the Advanced Computing Research Centre, University of Bristol -http://www.bris.ac.uk/acrc/.

All GWAS summary data used in this research, including that created for the applied examples, will be made publicly available.

The R scripts used in this manuscript, and GWASs created for it, are available at https://doi.org/10.17605/OSF.IO/YXZWC.

## Acknowledgements

We would like to thank Stephen Burgess and Jack Bowden for the feedback they provided on drafts of this manuscript.

## Conflicts of Interests & declarations

DG is employed part-time by Novo Nordisk. The remaining authors declare no conflicts of interest.

## References

1. Davies NM, Holmes MV, Smith GD. Reading Mendelian randomisation studies: a guide, glossary, and checklist for clinicians. BMJ. 2018 Jul 12;362:k601.

2. Burgess S, Davey Smith G, Davies NM, Dudbridge F, Gill D, Glymour MM, et al. Guidelines for performing Mendelian randomization investigations. Wellcome Open Res. 2020 Apr 28;4:186.

3. Hartwig FP, Davies NM, Hemani G, Smith GD. Two-sample Mendelian randomization: avoiding the downsides of a powerful, widely applicable but potentially fallible technique. International Journal of Epidemiology. 2016 Dec;45(6):1717–26.

4. single nucleotide polymorphism / SNP | Learn Science at Scitable [Internet]. [cited 2022 Apr 6]. Available from: http://www.nature.com/scitable/definition/single-nucleotide-polymorphism-snp-295

5. Lawlor DA. Commentary: Two-sample Mendelian randomization: opportunities and challenges. International Journal of Epidemiology. 2016 Jun;45(3):908–15.

6. Sadreev II, Elsworth BL, Mitchell RE, Paternoster L, Sanderson E, Davies NM, et al. Navigating sample overlap, winner’s curse and weak instrument bias in Mendelian randomization studies using the UK Biobank [Internet]. medRxiv; 2021 [cited 2022 Apr 6]. p. 2021.06.28.21259622. Available from: https://www.medrxiv.org/content/10.1101/2021.06.28.21259622v1

7. Woolf B, Di Cara N, Moreno-Stokoe C, Skrivankova V, Drax K, Higgins JPT, et al. Investigating the transparency of reporting in two-sample summary data Mendelian randomization studies using the MR-Base platform. International Journal of Epidemiology. 2022 Apr 6;dyac074.

8. PhD SPMF. Epidemiology for Canadian Students: Principles, Methods and Critical Appraisal. Brush Education; 2015. 304 p.

9. Davey Smith G, Hemani G. Mendelian randomization: genetic anchors for causal inference in epidemiological studies. Human Molecular Genetics. 2014 Sep 15;23(R1):R89–98.

10. Hernán MA, Robins JM. Causal Inference: What If.: 311.

11. Lash, L T, VanderWeele, J T, Haneuse, Sebastien, et al. Modern Epidemiology. 4th edition. Philadelphia: Lippincott Williams and Wilkins; 2021. 1192 p.

12. Keogh RH, Shaw PA, Gustafson P, Carroll RJ, Deffner V, Dodd KW, et al. STRATOS guidance document on measurement error and misclassification of variables in observational epidemiology: Part 1—Basic theory and simple methods of adjustment. Statistics in Medicine. 2020;39(16):2197–231.

13. van Smeden M, Lash TL, Groenwold RHH. Reflection on modern methods: five myths about measurement error in epidemiological research. International Journal of Epidemiology. 2020 Feb 1;49(1):338–47.

14. Chalak K. INSTRUMENTAL VARIABLES METHODS WITH HETEROGENEITY AND MISMEASURED INSTRUMENTS. Econometric Theory. 2017 Feb;33(1):69–104.

15. Jiang Z, Ding P. Measurement errors in the binary instrumental variable model. Biometrika. 2020 Mar 1;107(1):238–45.

16. Taylor AE, Davies NM, Ware JJ, VanderWeele T, Smith GD, Munafò MR. Mendelian randomization in health research: Using appropriate genetic variants and avoiding biased estimates. Economics & Human Biology. 2014 Mar 1;13:99–106.

17. Pierce BL, VanderWeele TJ. The effect of non-differential measurement error on bias, precision and power in Mendelian randomization studies. International Journal of Epidemiology. 2012 Oct 1;41(5):1383–93.

18. Dixon P, Hollingworth W, Harrison S, Davies NM, Davey Smith G. Mendelian Randomization analysis of the causal effect of adiposity on hospital costs. Journal of Health Economics. 2020 Mar 1;70:102300.

19. Arathimos R, Suderman M, Sharp GC, Burrows K, Granell R, Tilling K, et al. Epigenome-wide association study of asthma and wheeze in childhood and adolescence. Clinical Epigenetics. 2017 Dec;9(1):112.

20. Button KS, Ioannidis JPA, Mokrysz C, Nosek BA, Flint J, Robinson ESJ, et al. Power failure: why small sample size undermines the reliability of neuroscience. Nat Rev Neurosci. 2013 May;14(5):365–76.

21. Winner’s curse - Mendelian randomization dictionary [Internet]. [cited 2022 Apr 6]. Available from: https://mr-dictionary.mrcieu.ac.uk/term/winners-curse/

22. Barahona Ponce C, Scherer D, Brinster R, Boekstegers F, Marcelain K, Gárate-Calderón V, et al. Gallstones, Body Mass Index, C-Reactive Protein, and Gallbladder Cancer: Mendelian Randomization Analysis of Chilean and European Genotype Data. Hepatology. 2021;73(5):1783–96.

23. Morris TP, White IR, Crowther MJ. Using simulation studies to evaluate statistical methods. Statistics in Medicine. 2019;38(11):2074–102.

24. Sanderson E, Davey Smith G, Windmeijer F, Bowden J. An examination of multivariable Mendelian randomization in the single-sample and two-sample summary data settings. International Journal of Epidemiology. 2019 Jun 1;48(3):713–27.

25. Burgess S, Thompson SG. Multivariable Mendelian randomization: the use of pleiotropic genetic variants to estimate causal effects. Am J Epidemiol. 2015 Feb 15;181(4):251–60.

26. Carter AR, Sanderson E, Hammerton G, Richmond RC, Davey Smith G, Heron J, et al. Mendelian randomisation for mediation analysis: current methods and challenges for implementation. Eur J Epidemiol. 2021 May 1;36(5):465–78.

27. Yang Q, Sanderson E, Tilling K, Borges MC, Lawlor DA. Exploring and mitigating potential bias when genetic instrumental variables are associated with multiple non-exposure traits in Mendelian randomization [Internet]. medRxiv; 2019 [cited 2022 Apr 6]. p. 19009605. Available from: https://www.medrxiv.org/content/10.1101/19009605v1

28. Schooling CM, Lopez PM, Yang Z, Zhao JV, Au Yeung SL, Huang JV. Use of Multivariable Mendelian Randomization to Address Biases Due to Competing Risk Before Recruitment. Frontiers in Genetics [Internet]. 2021 [cited 2022 Apr 6];11. Available from: https://www.frontiersin.org/article/10.3389/fgene.2020.610852

29. Willer CJ, Schmidt EM, Sengupta S, Peloso GM, Gustafsson S, Kanoni S, et al. Discovery and Refinement of Loci Associated with Lipid Levels. Nat Genet. 2013 Nov;45(11):1274–83.

30. Scott RA, Lagou V, Welch RP, Wheeler E, Montasser ME, Luan J, et al. Large-scale association analyses identify new loci influencing glycemic traits and provide insight into the underlying biological pathways. Nat Genet. 2012 Sep;44(9):991–1005.

31. Woods SC, D’Alessio DA. Central Control of Body Weight and Appetite. J Clin Endocrinol Metab. 2008 Nov;93(11 Suppl 1):S37–50.

32. Trait: Glucose - IEU OpenGWAS project [Internet]. [cited 2022 Apr 6]. Available from: https://gwas.mrcieu.ac.uk/datasets/met-d-Glucose/

33. Richardson TG, Sanderson E, Palmer TM, Ala-Korpela M, Ference BA, Smith GD, et al. Evaluating the relationship between circulating lipoprotein lipids and apolipoproteins with risk of coronary heart disease: A multivariable Mendelian randomisation analysis. PLOS Medicine. 2020 Mar 23;17(3):e1003062.

34. Trait: Fasting glucose - IEU OpenGWAS project [Internet]. [cited 2022 Apr 6]. Available from: https://gwas.mrcieu.ac.uk/datasets/ieu-b-114/

35. Locke AE, Kahali B, Berndt SI, Justice AE, Pers TH, Day FR, et al. Genetic studies of body mass index yield new insights for obesity biology. Nature. 2015 Feb 12;518(7538):197–206.

36. FinnGen. FinnGen Documentation of R5 release. 2021; Available from: https://finngen.gitbook.io/documentation/v/r5/

37. Elsworth B, Lyon M, Alexander T, Liu Y, Matthews P, Hallett J, et al. The MRC IEU OpenGWAS data infrastructure [Internet]. bioRxiv; 2020 [cited 2022 Mar 30]. p. 2020.08.10.244293. Available from: https://www.biorxiv.org/content/10.1101/2020.08.10.244293v1

38. Zhao Q, Chen Y, Wang J, Small DS. Powerful three-sample genome-wide design and robust statistical inference in summary-data Mendelian randomization. Int J Epidemiol. 2019 Oct 1;48(5):1478–92.

39. Skrivankova VW, Richmond RC, Woolf BAR, Davies NM, Swanson SA, VanderWeele TJ, et al. Strengthening the reporting of observational studies in epidemiology using mendelian randomisation (STROBE-MR): explanation and elaboration. BMJ. 2021 Oct 26;375:n2233.

40. Turner S, Armstrong LL, Bradford Y, Carlson CS, Crawford DC, Crenshaw AT, et al. Quality Control Procedures for Genome-Wide Association Studies. Current Protocols in Human Genetics. 2011;68(1):1.19.1-1.19.18.

41. Marees AT, de Kluiver H, Stringer S, Vorspan F, Curis E, Marie-Claire C, et al. A tutorial on conducting genome-wide association studies: Quality control and statistical analysis. International Journal of Methods in Psychiatric Research. 2018;27(2):e1608.

42. Buniello A, MacArthur JAL, Cerezo M, Harris LW, Hayhurst J, Malangone C, et al. The NHGRI-EBI GWAS Catalog of published genome-wide association studies, targeted arrays and summary statistics 2019. Nucleic Acids Res. 2019 Jan 8;47(D1):D1005–12.

43. Fransson E, Knutsson A, Westerholm P, Alfredsson L. Indications of recall bias found in a retrospective study of physical activity and myocardial infarction. Journal of clinical epidemiology. 2008 Aug 1;61(8):840–7.

