## Supplementary Methods for "Re-evaluating the robustness of Mendelian randomisation to measurement error"

| Bias | 1) E->O* | 2) I->O* | 3) O->E* | 4) I->E* |
| --- | --- | --- | --- | --- |
| A) Instruments for the measurement error mechanism are pleiotropic and violate the exclusion restriction assumption. | 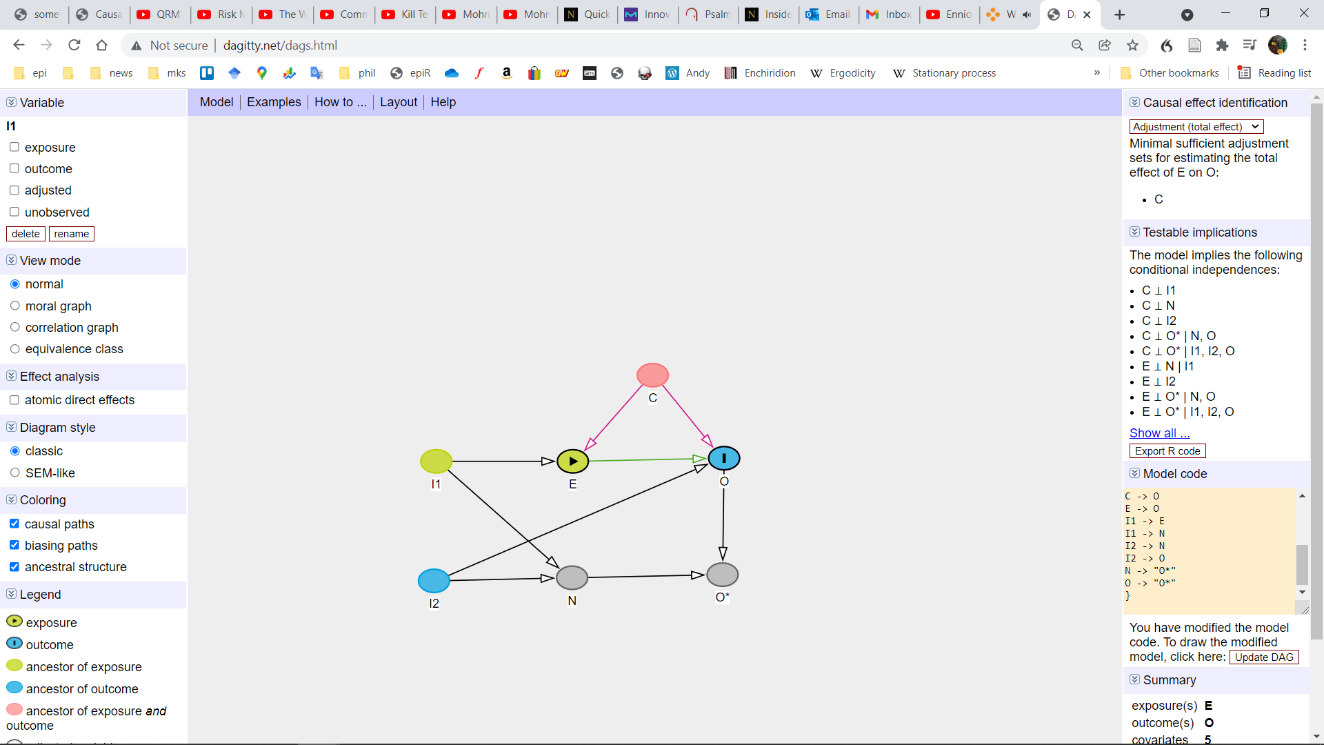 | 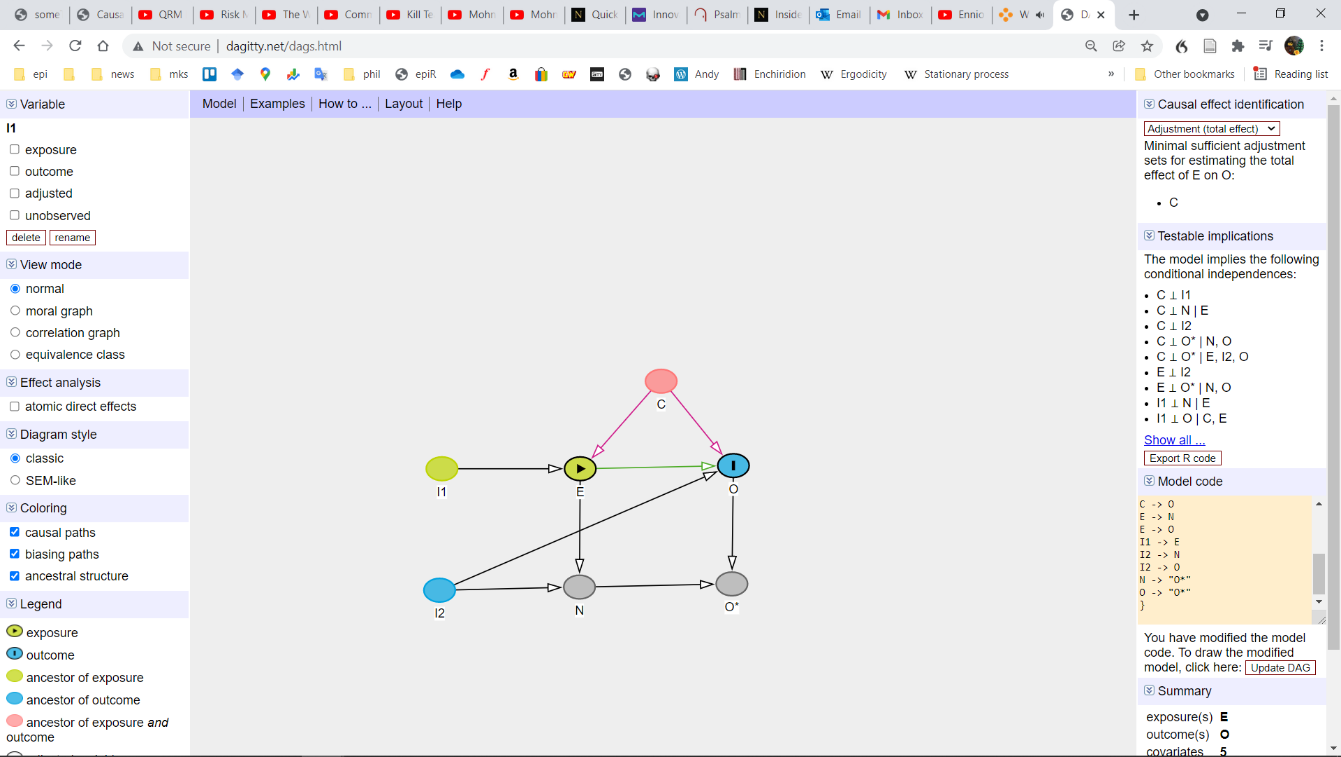 | 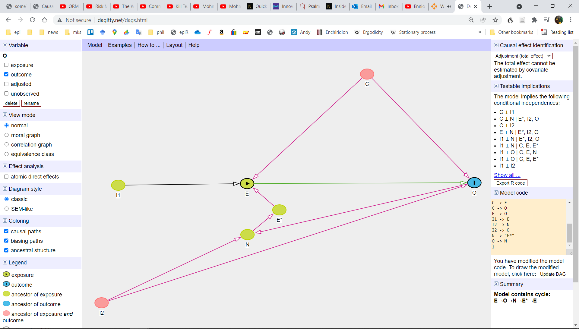 | 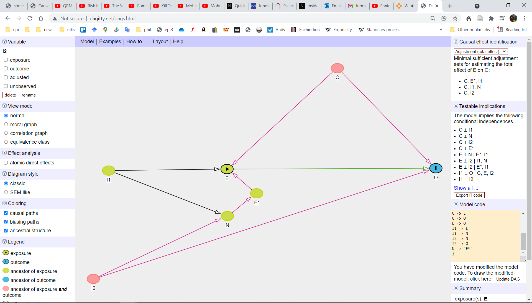 |
| B) The association between the instruments for the measurement error mechanism and the outcome is confounded (violating the independence assumption) | 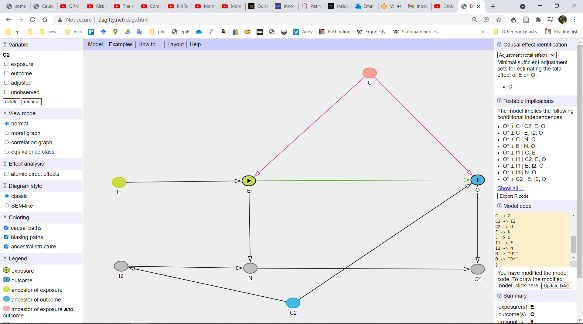 | 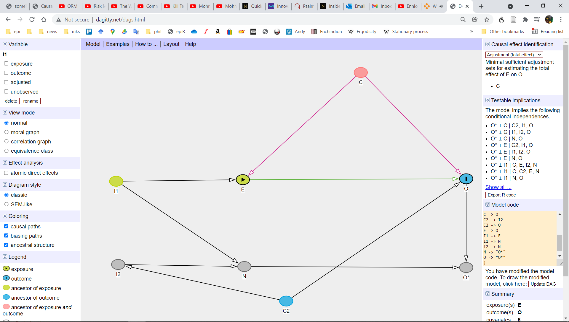 | 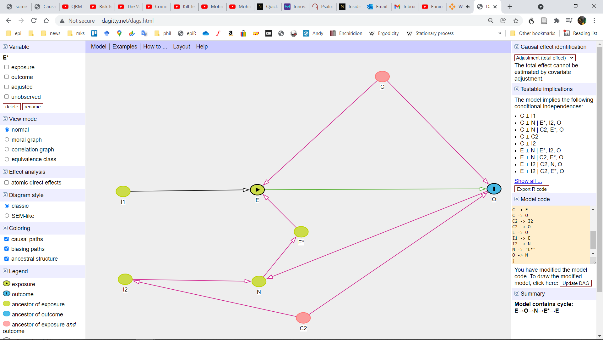 | 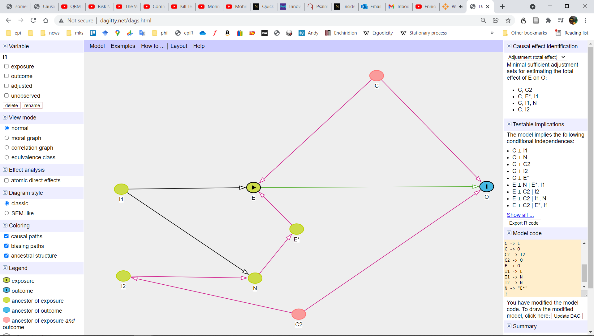 |
| C) Instruments for the measurement error mechanism are weak. | 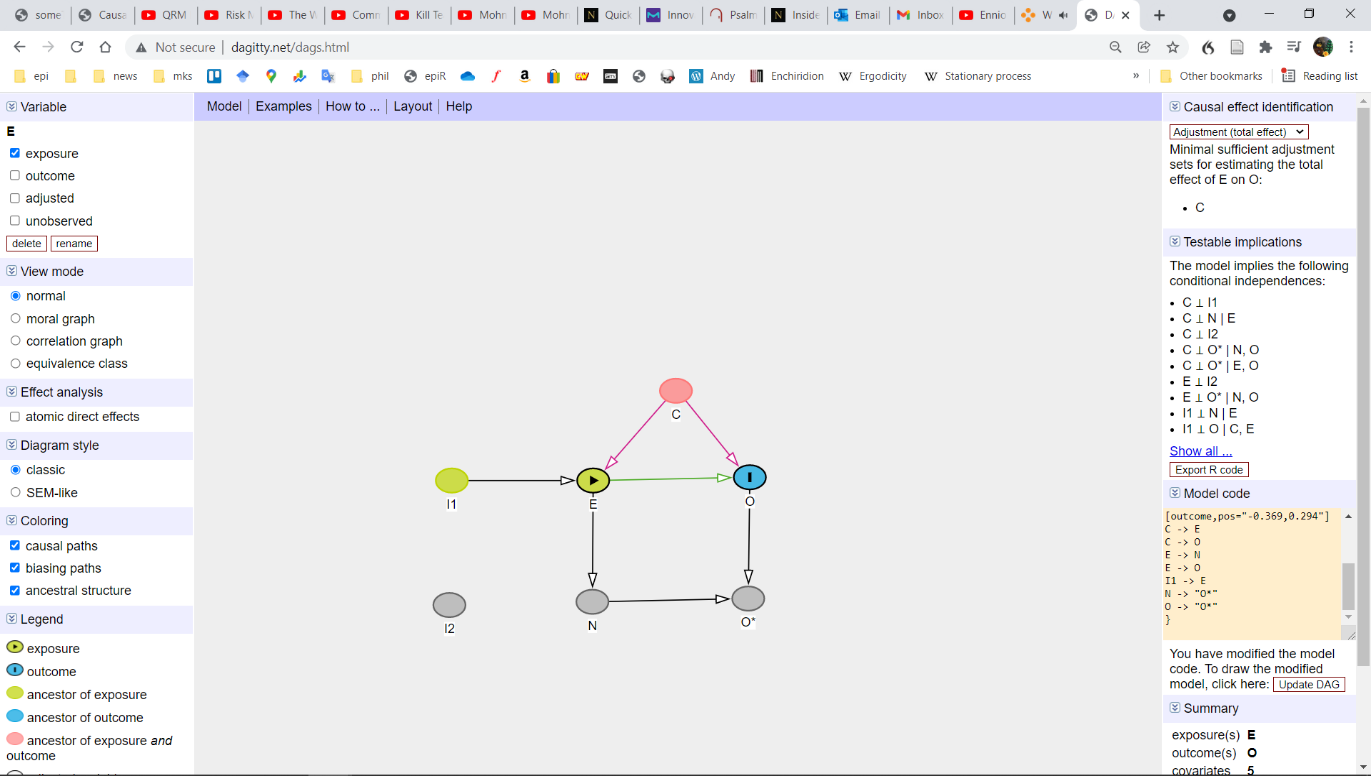 | 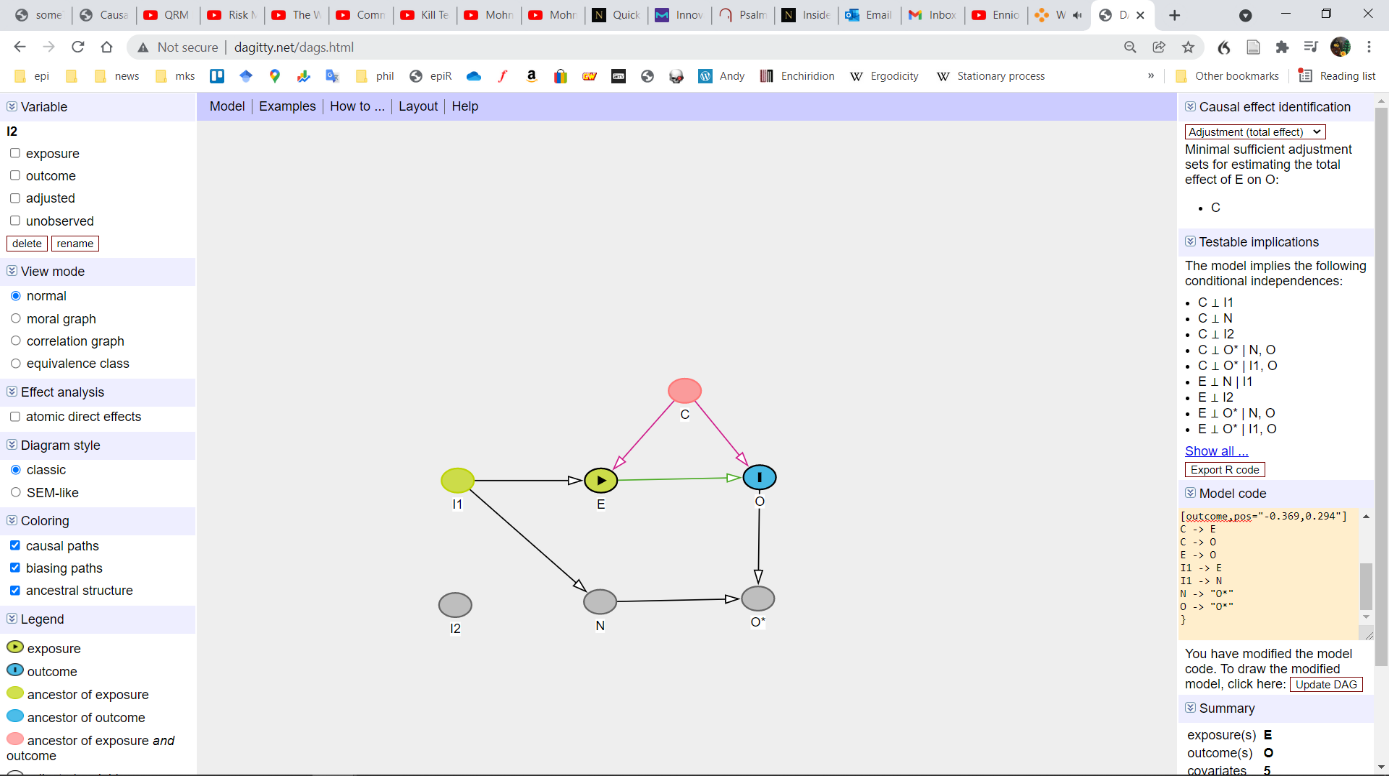 | 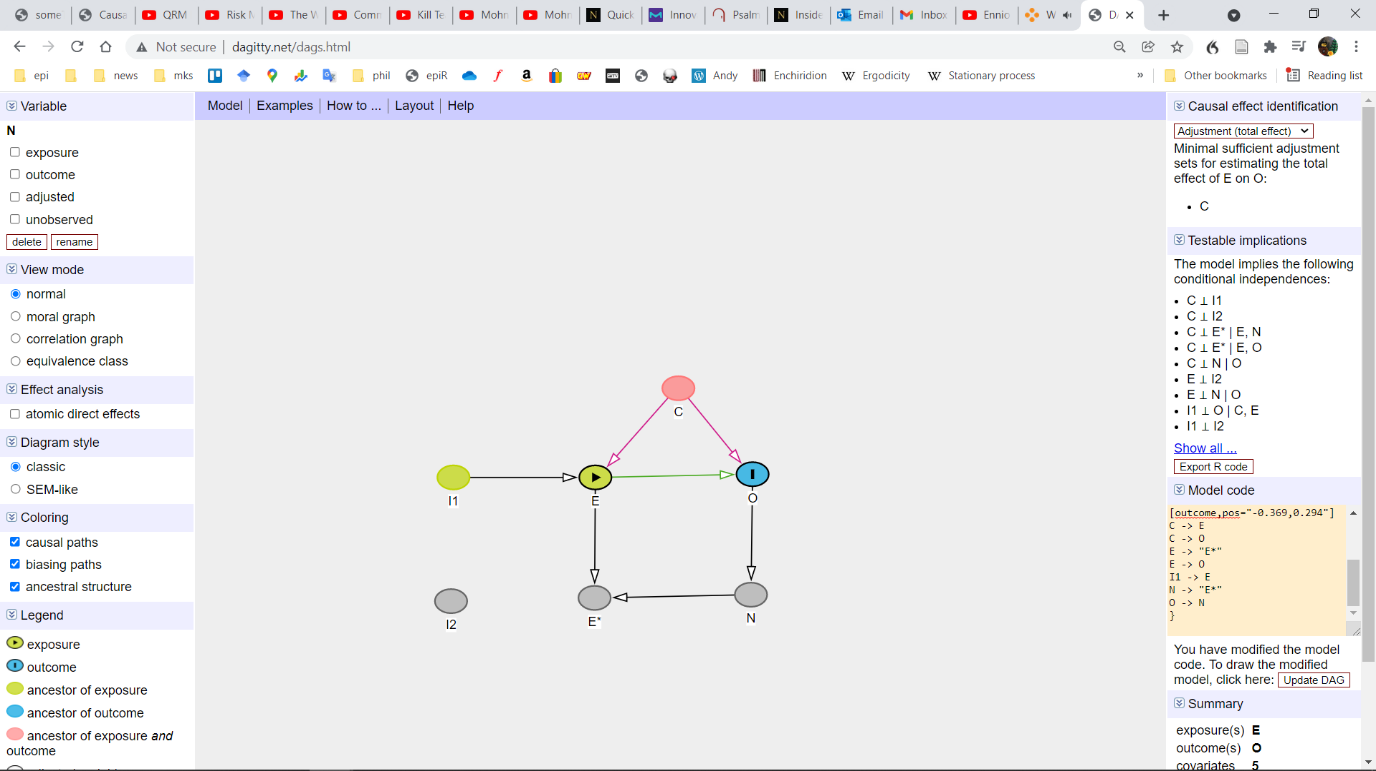 | 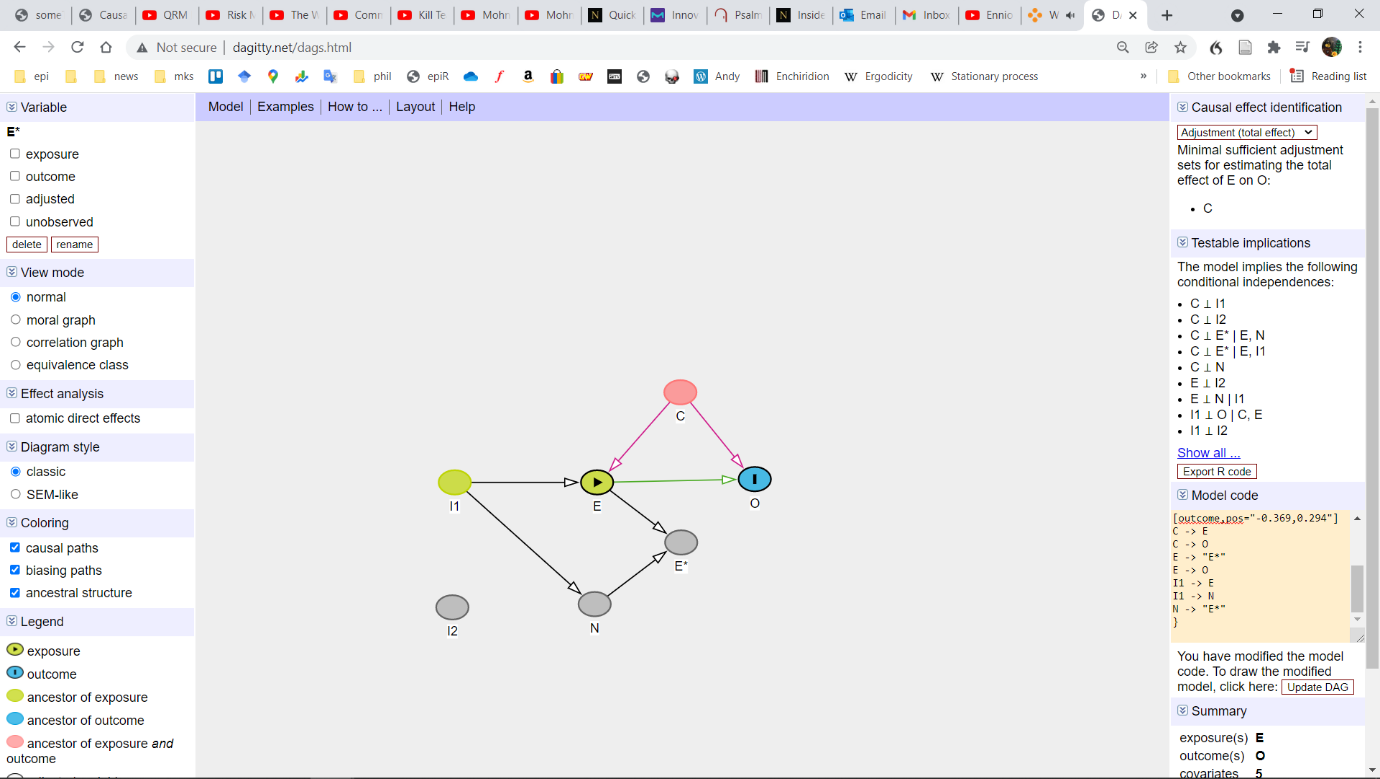 |
| D) Imperfect measure of cause of differential measurement error in outcome/exposure (using an r of 0.7 and 0.8). | 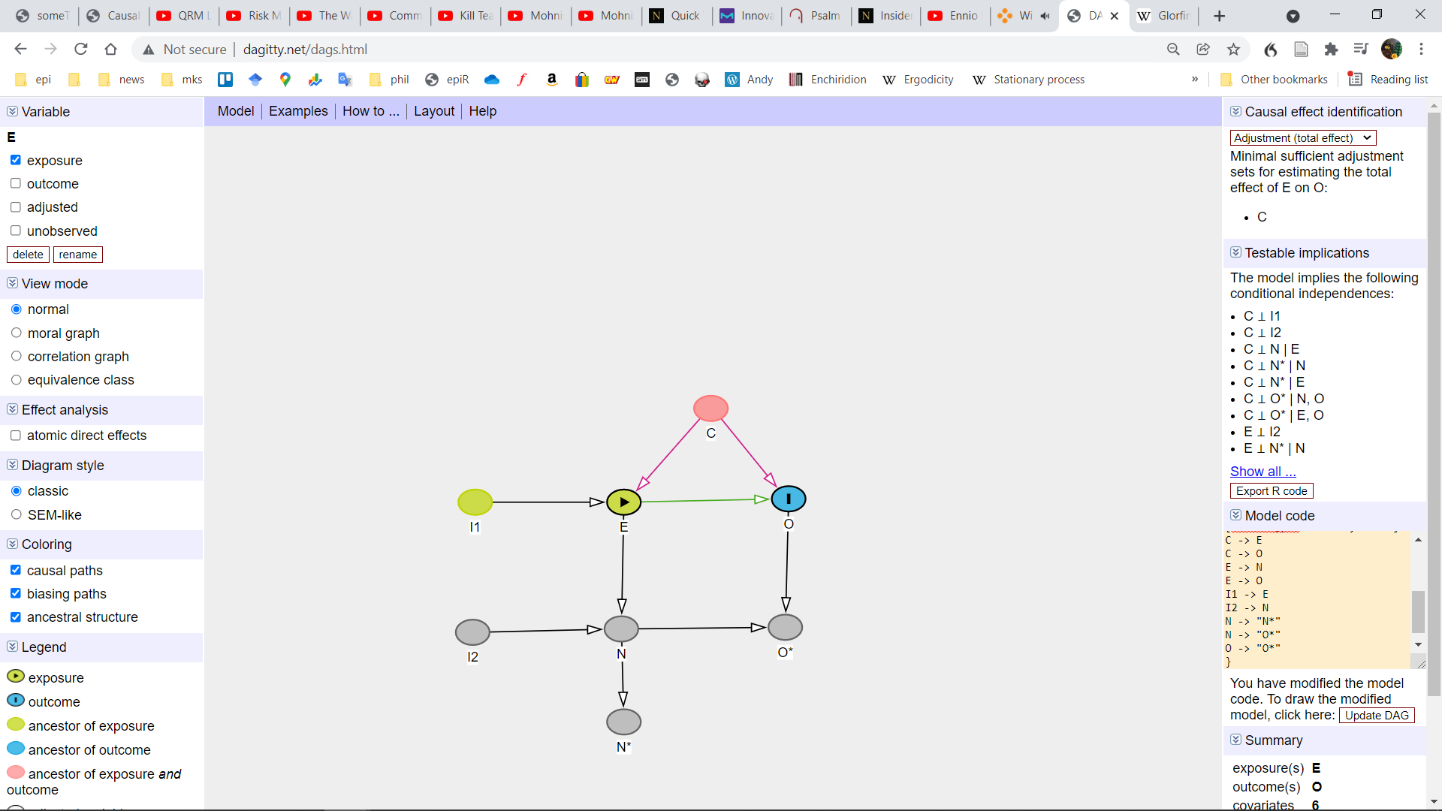 | 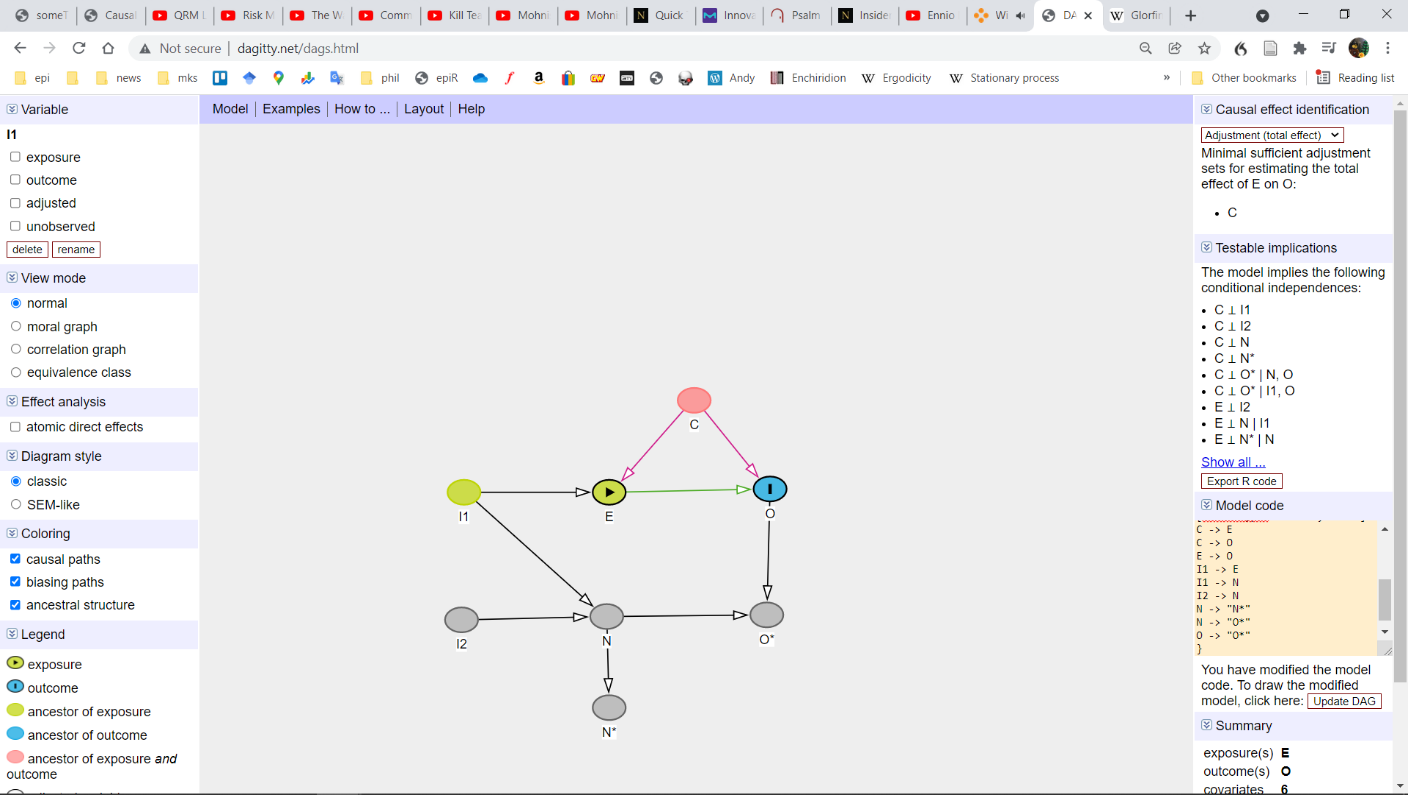 | 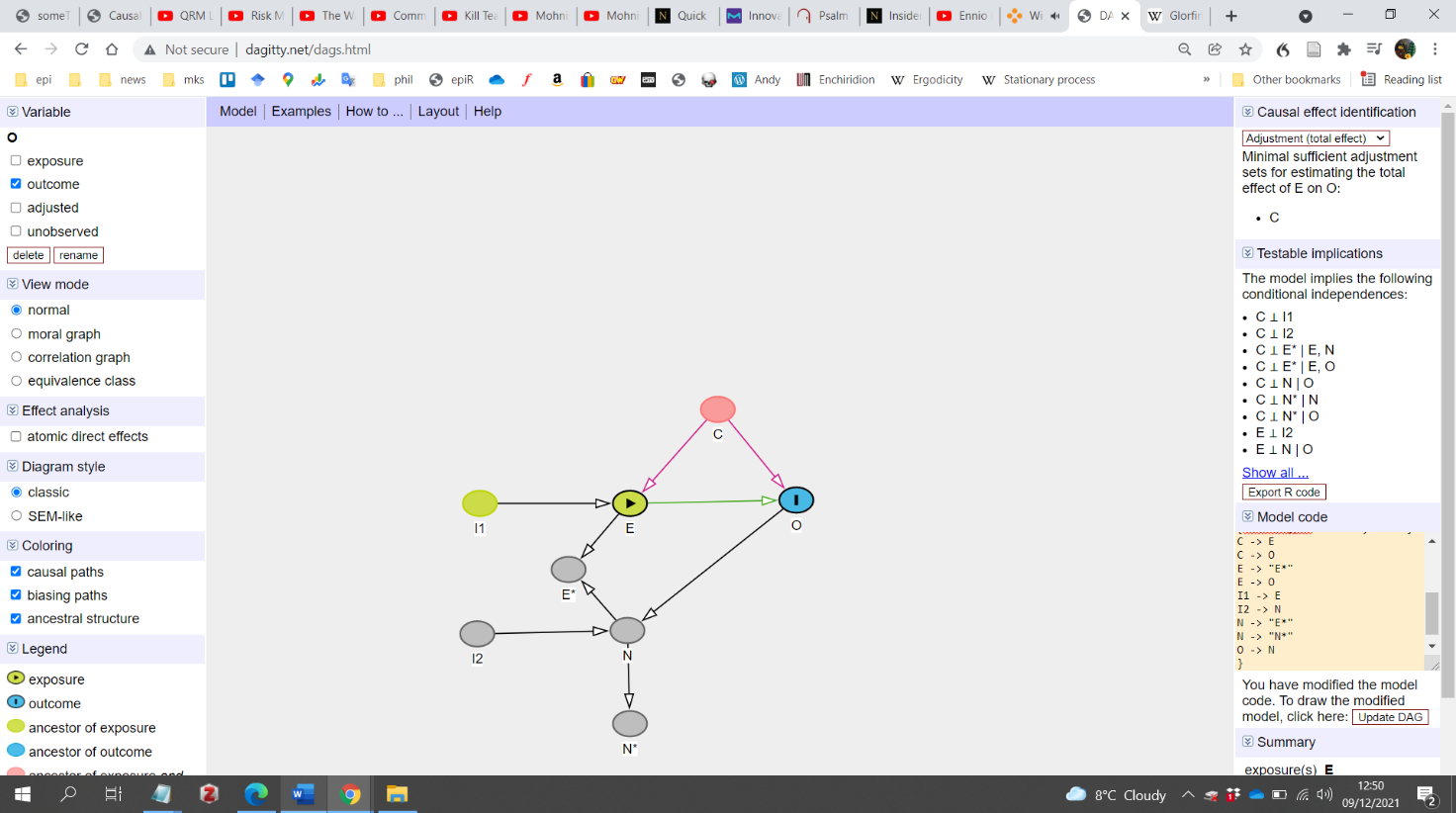 | 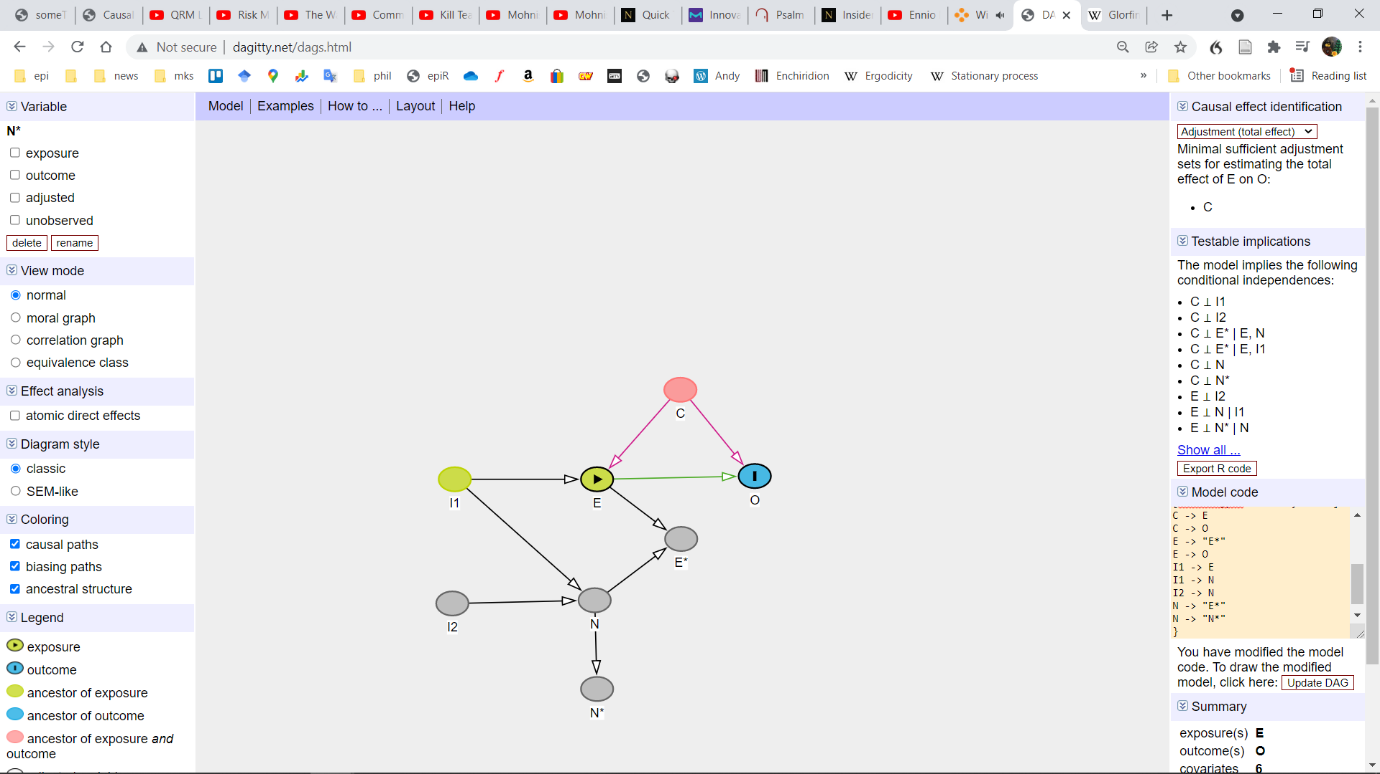 |
| E) The measurement error mechanism is only partially mediated by N | 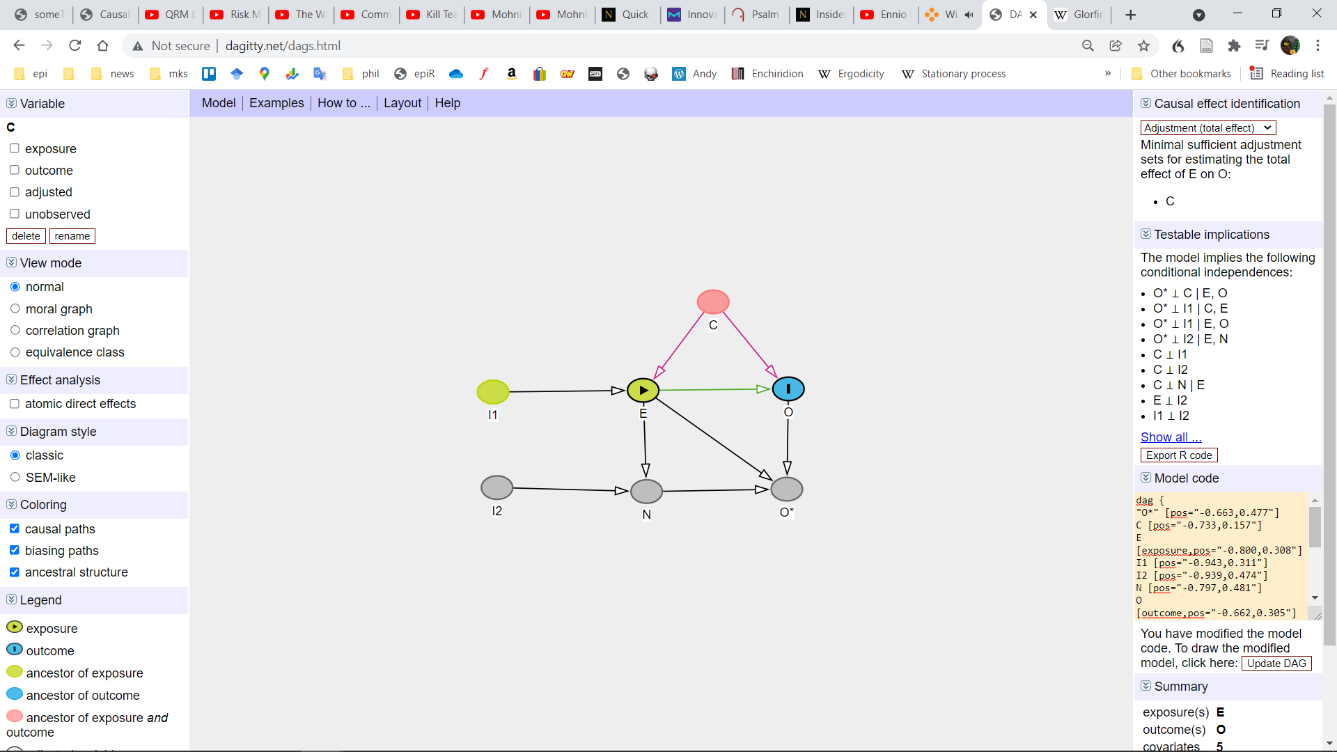 | 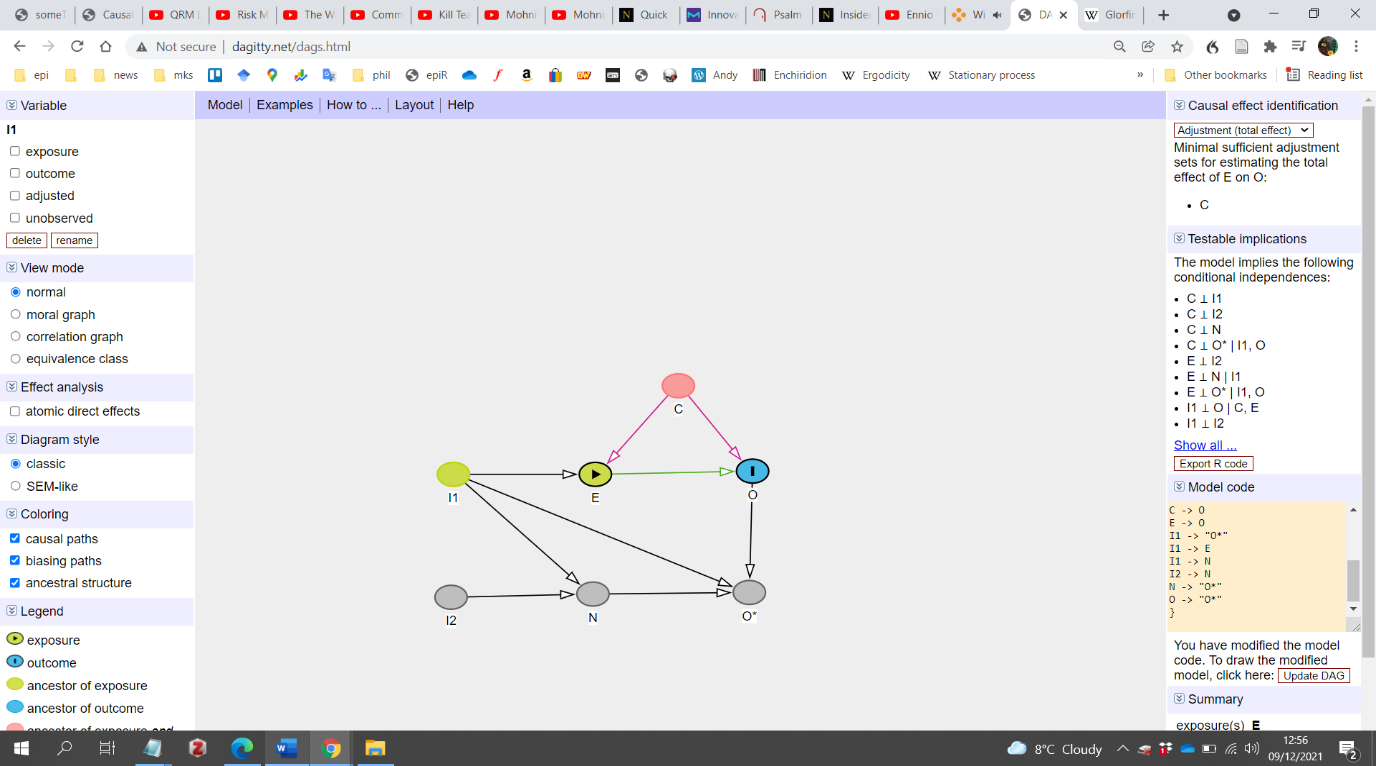 | 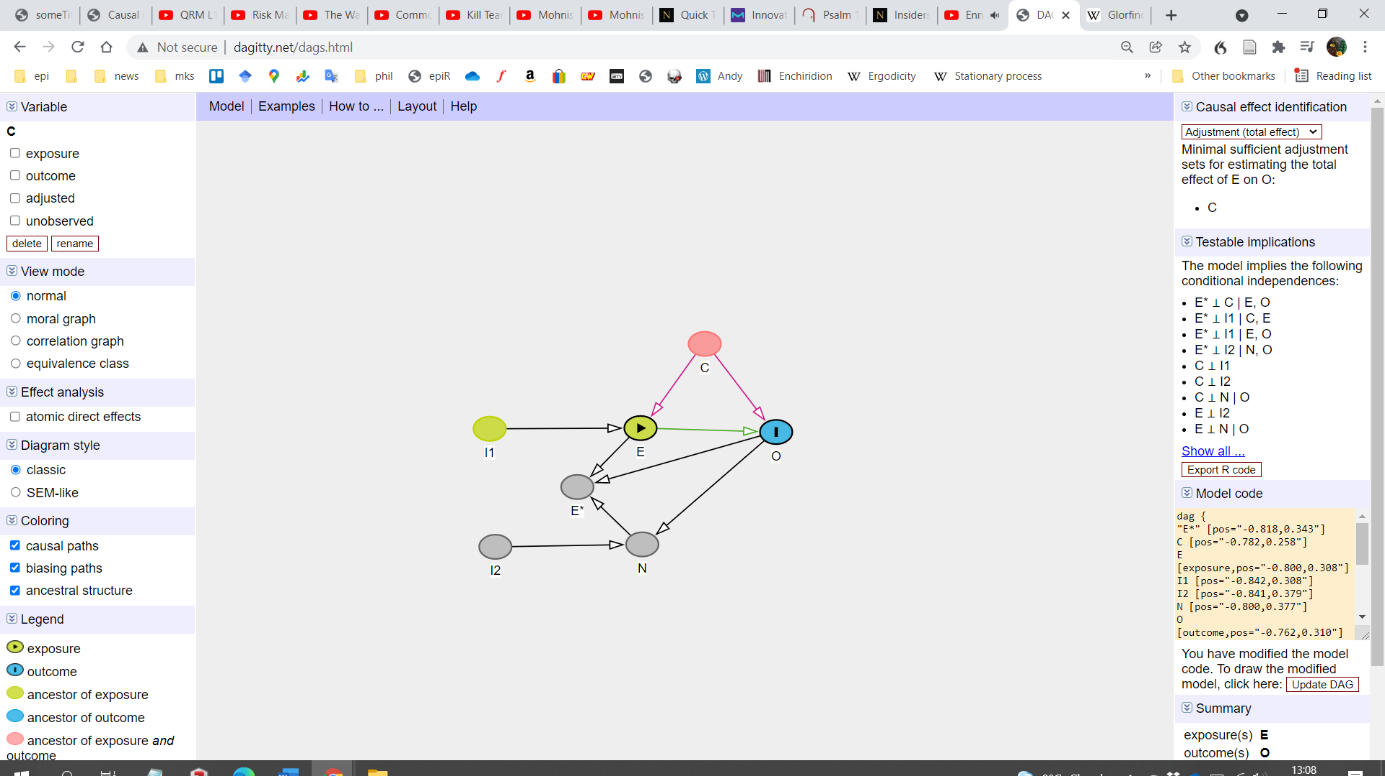 | 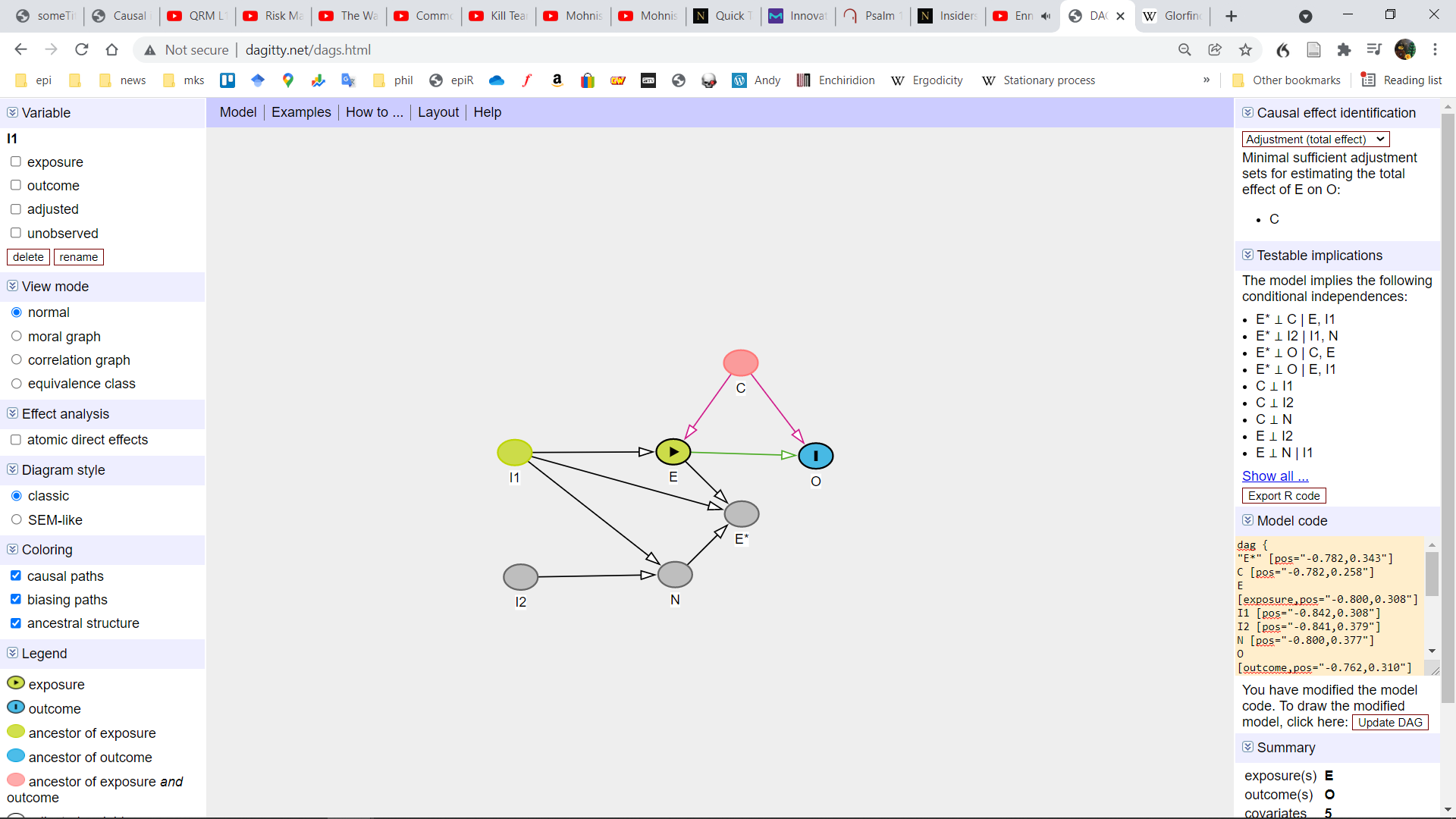 |
| F) The measurement error mechanism is a confounder rather than a mediator. | 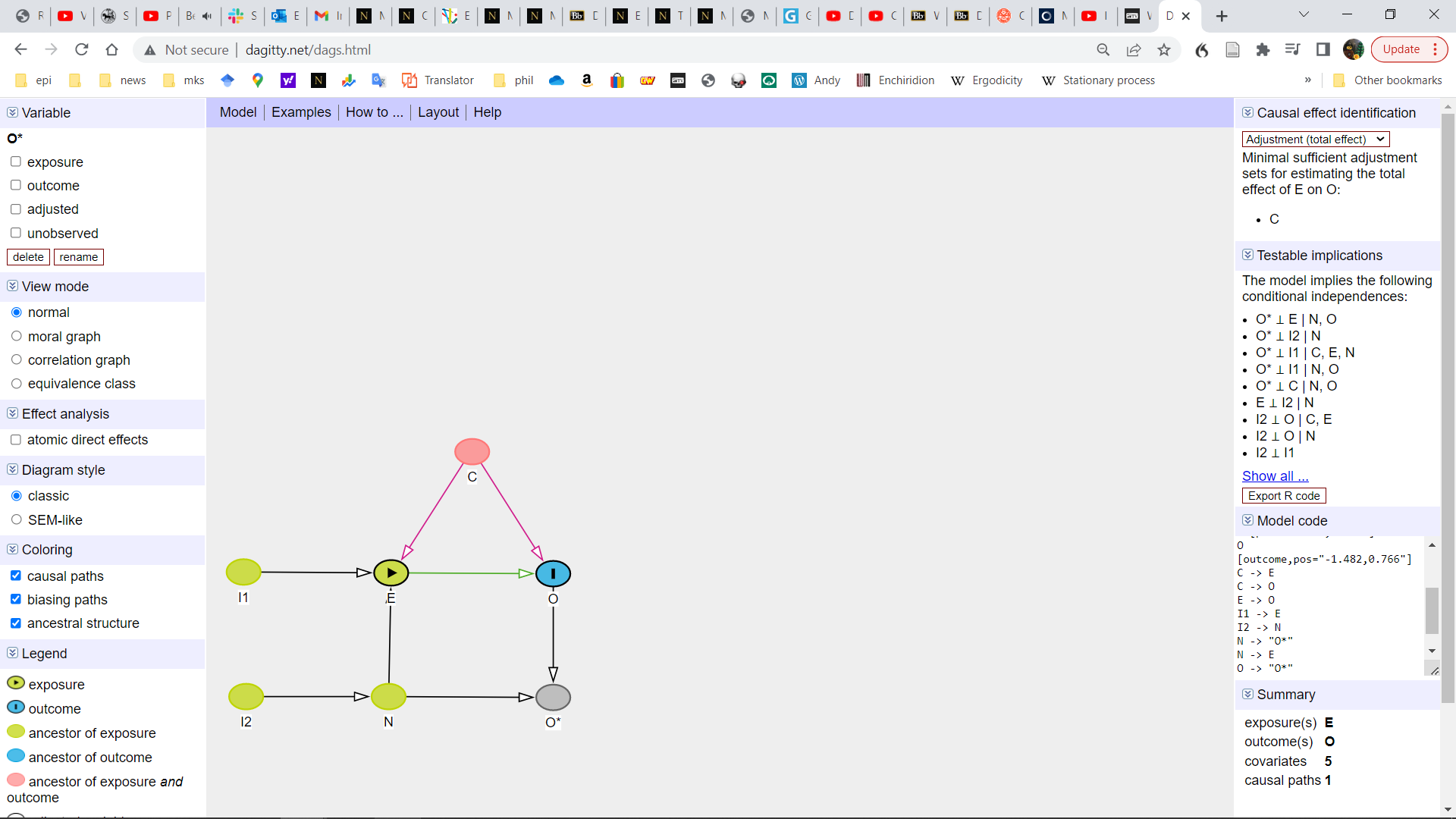 | 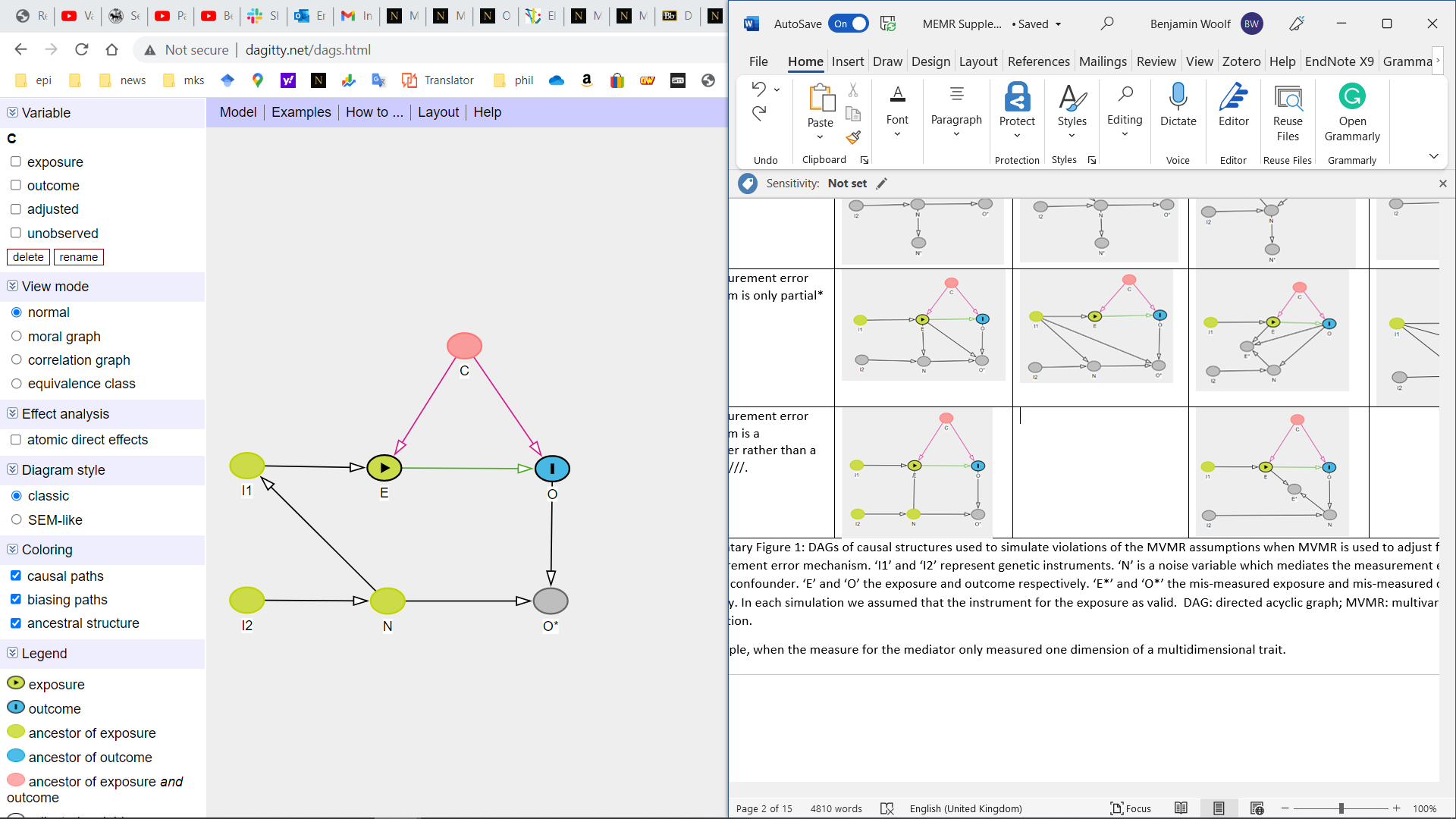 | 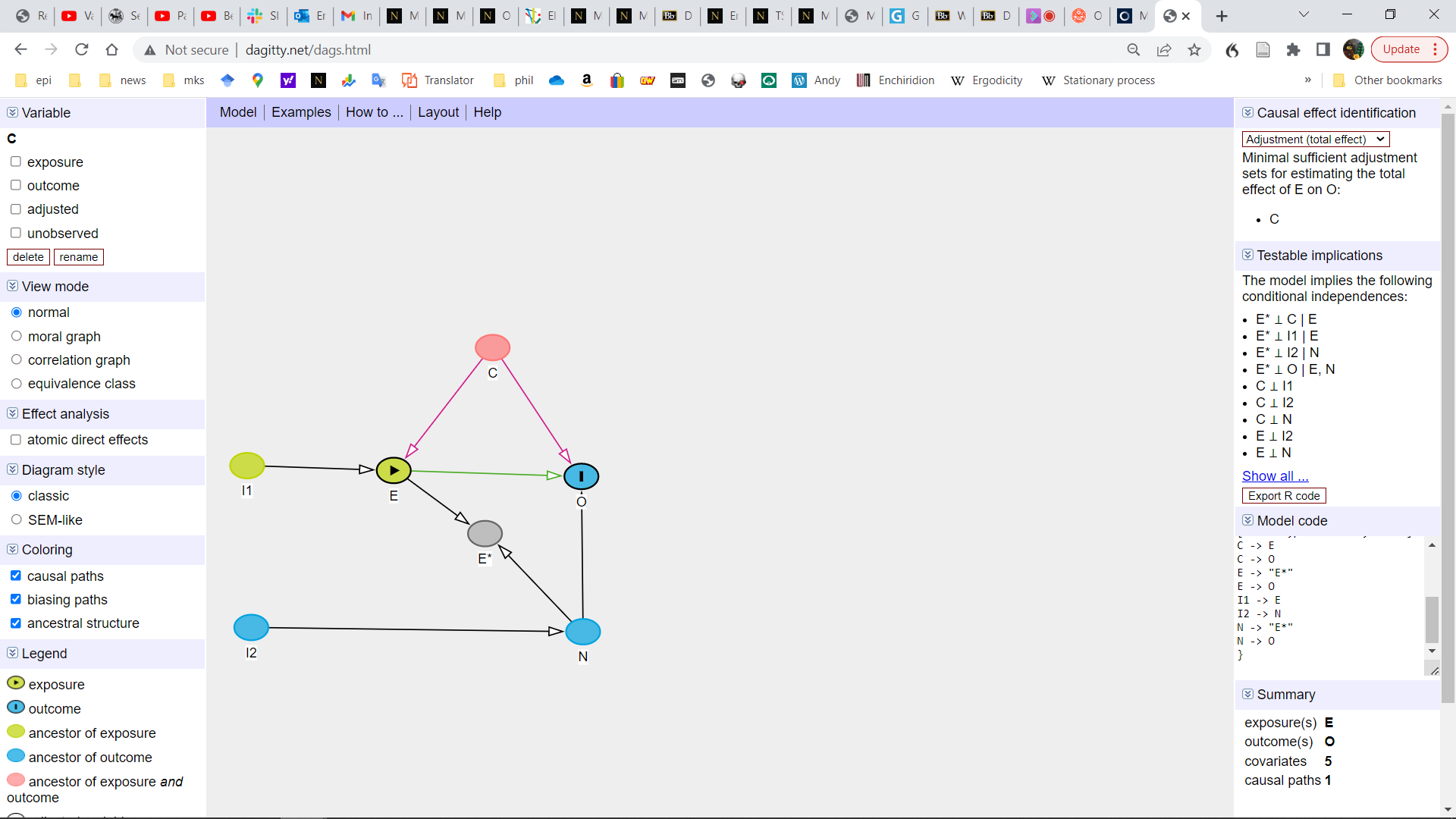 | 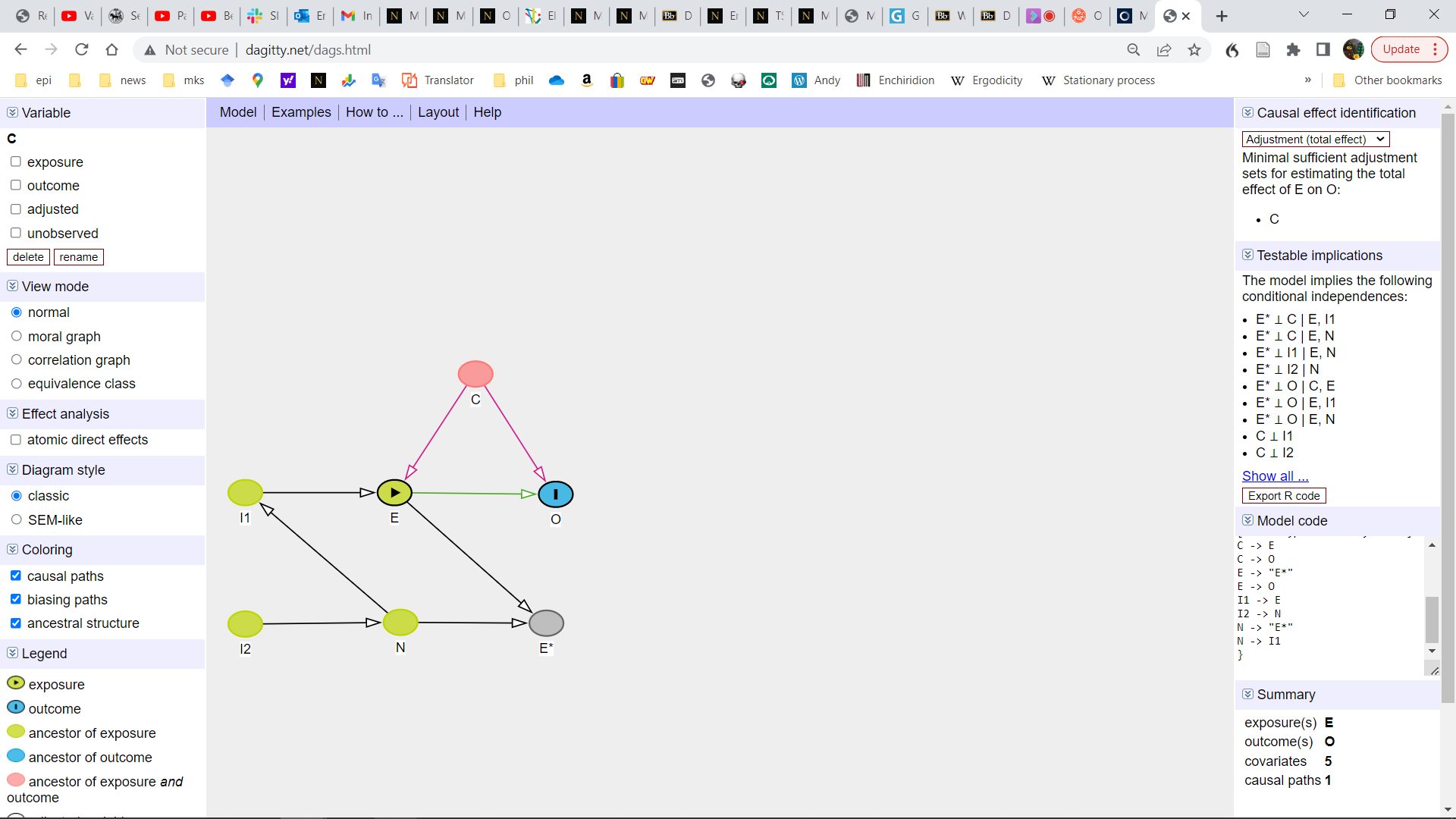 |

Supplementary Figure 1: DAGs of causal structures used to simulate violations of the MVMR assumptions when MVMR is used to adjust for a mediator of the measurement error mechanism. ‘I1’ and ‘I2’ represent genetic instruments. ‘N’ is a noise variable which mediates the measurement error mechanism, and ‘C’ is a confounder. ‘E’ and ‘O’ the exposure and outcome respectively. ‘E*’ and ‘O*’ the mis-measured exposure and mis-measured outcome respectively. In each simulation we assumed that the instrument for the exposure as valid. DAG: directed acyclic graph; MVMR: multivariable Mendelian randomization.

* For example, when the measure for the mediator only measured one dimension of a multidimensional trait.

**Supplementary Methods**

**1. Simulation study to explore the effect of differential measurement error in the exposure or outcome on MR estimates**

We report our simulations using the ADEMP (aims, data-generating mechanisms, estimands, methods, and performance measures) approach:(1)

*Aims*: The aim of this simulation was, as a simple proof of concept, to explore if estimates from MR models are affected by the presence of the types of differential measurement error defined in the main text. As a comparison, we also look at the effect of the bias on the linear regression estimate that would be used in a traditional epidemiological association study. Genetic correlations are a related method to MR, which can be used to estimate the extent to which two traits have a shared genetic overlap. Because of this, we further explore the effect of differential measurement error on the genetic correlation between the two traits, and benchmark it with the phenotypic correlation between the two traits.

*Data-generating mechanisms*: The “DAG without mediator of the measurement error mechanism” column of Table 1 presents four DAGs, based on those in Hernan and Robins’,(2) which represent these differential measurement errors. These were used as the basis for the data generation mechanism for the simulation. Each DAG’s nodes were simulated as standard normal variables (mean = 0, standard deviation [SD] = 1). Each arrow was used to represent a linear causal effect of the node from which the arrow originated on the node to which the arrow was directed, with each beta value being set as 1. The two-sample MR simulation simulated two samples each with 500,000 participants, and the 1SMR, observational, and correlational analyses uses one 500,000 person sample. The simulations were each repeated 1000 times.

More formally, we simulated the genetic liability to the exposure I_1_ and the confounder as standard Normal (0,1) random variables:

I_1_ ~ N(0,1)

C ~ N(0, 1)

The true (error-free) exposure was then simulated as:

E = I_1_ + C + ε_1_

where ε_1_ ~ N(0, 1).

The genetic liability to the outcome was simulated as a Normal random variable:

I_2_ ~ N(0, 1)

The true (error-free) outcome was then simulated as:

O = β_0_*E + C + I_2_ + ε_2_

where ε_2_ ~ N(0, 1), and β_0_=0 where there is in truth no causal effect and β_0_=1 otherwise

We simulated systematic outcome-related measurement error in the exposure (scenario A1, Table 1) as:

M_E_ = O + ε_5_

where ε_5_ ~ N(0, 1).

This gives an observed exposure of:

E*_o_ = E + M_E_

We simulated systematic exposure -related measurement error in the outcome (scenario B1, Table 1) as:

M_O_ = E+ ε_6_

where ε_6_ ~ N(0, 1).

This gives an observed outcome of:

O*_E_ = O + M_O_

We simulated systematic instrument-related measurement error in the exposure (scenario C1, Table 1) as:

M_IE_ = I_1_ + ε_8_

where ε_8_ ~ N(0, 1).

This gives an observed exposure of:

E*_I_ = E + M_IE_

We simulated systematic instrument-related measurement error in the outcome (scenario D1, Table 1) as:

M_IO_ = I_1_ + ε_9_

where ε_9_ ~ N(0, 1).

This gives an observed outcome of:

O*_I_ = O +	M_IO_

*Estimands and other targets*: The estimand was the average causal effect of intervening and increasing the exposure of each individual from its observed level x by a single unit, or E[Y(X=x+1)] – E[Y(X=x)] using the Potential Outcomes framework.

*Methods*: We compare five methods of estimating the exposure-outcome association: 1) the two-sample MR Wald ratio was calculated using the code from the mr_wald function in the TwoSampleMR R package. 2) The two-stage least square regression for the 1SMR estimator was calculated using the ivreg function from the eponymous R package.(3,4) 3) a linear regression that adjusted for all confounding. Although it is unrealistic for an observational study to adjust for all confounders in practice, by doing so here, we can show the exclusive impact of measurement error on these estimates. 4) The genetic correlation between exposure and outcome, calculated as the correlation between the weighted genetic risk score (GRS) for the exposure and the weighted GRS for the outcome. The GRS for the (mis-measured) exposure was created from the values of the exposure predicted by its genetic instrument in a linear regression model. The GRS for the (mis-measured) outcome was created from the values of the outcome predicted by both its own unique genetic instrument and the genetic instrument for the exposure (to account for the mediated pleiotropic effect of the exposure’s GRS on the outcome). 5) The correlation between the exposure and the outcome.

*Performance measure:* The performance measure was the bias and 95% confidence interval coverage for these five methods. Bias is defined as the difference between the value of the estimate that was observed and the true value. The coverage of the 95% confidence interval is defined as the proportion of times that the 95% confidence interval for the estimate includes the true estimate. Because the true value of the genetic correlation under the alternative was not defined *a priori* it was estimated ‘empirically’ by simulating it without any bias. However, all other ‘true’ values were defined *a priori*, and this number was used in the calculation of bias.

**2. Simulation study to explore the use of multivariable MR to reduce bias due to differential measurement error in the exposure or outcome.**

*Aim*: The aim of the second simulation was to explore if MVMR can be used to attenuate non-differential measurement error, and, then if it can do so under violations of its identifying assumptions.

*Data-generating mechanisms*: We first simulated the DAGs in “DAGs with mediator of the measurement error mechanism” column of Table 1 as the data generation mechanism for exploring the use of MVMR to attenuate bias from differential measurement error. These four DAGs are similar to those presented in the previous column, however, they include a mediator of the measurement error mechanism, and a genetic instrument for the measurement error mechanism. We then simulated the DAGs in Supplementary Figure 1 to explore the effect of threats to the validity of the instrument for the measurement error mechanism on the ability of MVMR to attenuate bias. Specifically, we explored what would happen when either, there was weak instrument bias for the genetic instrument for the measurement error mechanism, the genetic instrument for the measurement error mechanism exerted a pleiotropic effect on the outcome, there was a residual confounder of the genetic instrument for the measurement error mechanism and the outcome, the measurement error mechanism only partially mediates the differential error, in which the measurement error mechanism was a confounder rather than a mediator, or finally, the measurement error mechanism was measured through a non-differentially miss-measured proxy with a correlation of 0.7 or 0.8. As before, all nodes in the DAG were simulated as random normal variables (mean of 0 and an SD of 1) with a sample size of 500,000; and all betas are 1. The exception to this was for the simulation where the measurement error mechanism only partially mediates the path on which the differential error occurs. In this instance, the direct effect from both mediator and the biasing variable on the variable being biased was 0.5, so the total effect of the bias was 1. Classical measurement error in the measure of the measurement error mechanism was simulated so that the correlation between the measurement error mechanism and its error term were approximately 0.7 and 0.8 by modelling the SD of these nodes as the SD of the measurement error mechanism multiplied by 1.01 and 0.75 respectively. Each simulation was repeated 1000 times.

More formally, we simulated the genetic liability to the exposure as a standard Normal random variable:

I_1_ ~ N(0,1)

Likewise, we simulated the genetic liability to the measurement error mechanism as an independent standard Normal random variable:

I_2_ ~ N(0,1)

The confounder of the exposure-outcome association was simulated as a standard Normal random variable:

C_1_ ~ N(0, 1)

The true exposure was then simulated as:

E = I_1_ + C_1_ + ε_1_

where ε_1_ ~ N(0, 1).

The true outcome was then simulated as:

O = E + C_1_ + ε_2_

where ε_2_ ~ N(0, 1).

We simulated a pleiotropic effect between the genetic liability to the measurement error mechanism and the outcome (scenarios from A rows in Supplementary Figure 1) using the following formula:

O_p_ = E + C_1_ + I_2_ + ε_2_

where ε_2_ ~ N(0, 1).

We simulated population structure as a random normal variable

PS ~ N(0, 1)

And then simulated confounding between the instruments for the measurement error mechanism and the outcome (scenarios from B rows in Supplementary Figure 1) by adding it as a confounder of the instrument (for the measurement error mechanism) and the true outcome association:

I_2,ps_ = I_2_ + PS

O_ps_ = O + PS

We simulated a weak instrument (scenarios from C rows in Supplementary Figure 1) by re-defining the instrument as an identically distributed normal variable which was not associated with the exposure:

I_2,wi_ ~ N(0, 1)

When there was exposure-related measurement error in the outcome (scenarios in column 1 of Supplementary Figure 1), we simulated the measurement error mechanism as:

N_e->o_ = E + I_2_ + ε_3_

where ε_3_ ~ N(0, 1).

The observed outcome (with exposure-related measurement error) was then simulated as

O’_e->o_ = O - N_e->o_ + ε_4_

where ε_4_ ~ N(0, 1). N_e->o_ was simulated as subtractive, rather than additive, with O here so that the bias was positive rather than negative.

We simulated a setting where the covariate included in the MVMR model was an imperfect measure of the different measurement error mechanism by adding classical (non-differential) measurement error (scenario 1D in Supplementary Figure 1) to the measure of the (differential) measurement error mechanism included in the MVMR model:

N’_0.8,e->o_ = N_e->o_ + N(0, [0.75*SD(N_e->o_)]^2^)

N’_0.7,e->o_ = N_e->o_ + N(0, [1.01*SD(N_e->o_)]^2^)

Where SD(N_e->o_) is the standard deviation of the M_e->o_ term. The multiple of 0.75 and 1.01 were to create an approximate correlation between the mechanism and its error prone measure of 0.8 and 0.7 respectively.

To simulate pleiotropy in this setting (scenario 1A in Supplementary Figure 1) we then added a direct effect of the genetic liability for the measurement error mechanism and the measurement error term

O’_e->o, p_ = O_p_ + N_e->o_ + ε_5_

where ε_5_ ~ N(0, 1).

To simulate population structure (scenario 1B in Supplementary Figure 1) in this setting we modelled measurement error mediator as:

N_e->o,ps_ = E + I_2,ps_ + ε_6_

where ε_6_ ~ N(0, 1).

And the mismeasured outcome as:

O’_ps, e->o_ = O_ps_ - N_e->o,ps_ + ε_7_

where ε_7_ ~ N(0, 1). N_e->o,ps_ was simulated as subtractive, rather than additive, with O here so that the bias was positive rather than negative.

To simulate the setting where we have only partially measured the mechanism through which the measurement error occurs (i.e. the measurement error is only partially mediated by the variable being adjusted for) (scenario 1E in Supplementary Figure 1) we simulated the differential error in outcome as a function of both the mediator and a direct effect of the exposure:

O’_part, e->o_ = O - N_e->o_/2 – E/2 + ε_8_

where ε_8_ ~ N(0, 1). E was simulated as having a negative effect so that the residual bias was positive rather than negative.

When there was outcome related measurement error in the exposure (scenarios in column 3 in Supplementary Figure 1), we simulated the measurement error mechanism as:

N_o->e_ = O + I_2_ + ε_9_

where ε_9_ ~ N(0, 1).

The observed, miss measured, exposure was then simulated as:

E’_o->e_ = N_o->e_ + E + ε_10_

where ε_10_ ~ N(0, 1).

We simulated a setting where the covariate included in the MVMR model was an imperfect measure of the different measurement error mechanism by adding classical (non-differential) measurement error (scenario 3D in Supplementary Figure 1) to the measure of the (differential) measurement error mechanism included in the MVMR model:

N’_0.8,o->e_ = N_o->e_ + N(0, [0.75*SD(N_o->e_)]^2^)

N’_0.7,o->e_ = N_o->e_ + N(0, [1.01*SD(N_o->e_)]^2^)

Where SD(N_o->e_) is the standard deviation of the N_o->e_ term. The multiple of 0.75 and 1.01 were to create an approximate correlation between the mechanism and its error prone measure of 0.8 and 0.7 respectively.

To simulate pleiotropy (scenario 3A in Supplementary Figure 1) in this setting we then added a direct effect of the genetic liability for the measurement error mechanism we first re-simulated the term for the mechanism:

O_o->e, p_ = O_p_ + N_o->e_ + ε_11_

where ε_11_ ~ N(0, 1).

We then simulated the error prone, miss measured, exposure as:

E’_o->e,p_ = N_o->e,p_ + E + ε_12_

where ε_12_ ~ N(0, 1^2^).

To simulate population structure (scenario 3B in Supplementary Figure 1) in this setting we modelled the measurement error mediator as:

N_o->e,ps_ = O_ps_ + I_2,ps_ + ε_13_

where ε_13_ ~ N(0, 1^2^).

And the observed, error prone, exposure as:

E’_o->e,ps_ = E + N_o->e,ps_ + ε_14_

where ε_14_ ~ N(0, 1^2^).

To simulate the setting where we have only partially measured the mechanism through which the measurement error occurs (i.e. the measurement error is only partially mediated by the variable being adjusted for) (scenario 3E in Supplementary Figure 1) we simulated the differential error in the exposure as a function of both the mediator and a direct effect of the outcome:

E’_part, o->e_ = E – N_o->e_/2 – O/2 + ε_15_

where ε_15_ ~ N(0, 1^2^). O was simulated as having a negative effect so that the residual bias was positive rather than negative.

We simulated the measurement error mechanism when there was instrument related measurement error (Scenarios in columns 2 and 4 in Supplementary Figure 1) as:

N_i_ = I_1_ + I_2_ + ε_16_

where ε_16_ ~ N(0, 1^2^).

We simulated a setting where the covariate included in the MVMR model was an imperfect measure of the different measurement error mechanism by adding classical (non-differential) measurement error (scenarios 2D and 4D in Supplementary Figure 1) to the measure of the (differential) measurement error mechanism included in the MVMR model:

N’_0.8,I_ = N_I_ + N(0, [0.75*SD(N_I_)]^2^)

N’_0.7,I_ = N_I_ + N(0, [1.01*SD(N_I_)]^2^)

Where SD(N_I_) is the standard deviation of the N_I_. The multiple of 0.75 and 1.01 were to create an approximate correlation between the mechanism and its error prone measure of 0.8 and 0.7 respectively.

When there was instrument related measurement error in the exposure (Scenarios in column 4 in Supplementary Figure 1), we simulated the observed, error prone, exposure as:

E’_i_ = E + N_i_ + ε_17_

where ε_17_ ~ N(0, 1^2^)

When there was instrument related measurement error in the exposure (Scenarios in column 3 in Supplementary Figure 1), we simulated the observed, error prone, outcome as:

O’_i_ = O + N_i_ + ε_18_

where ε_18_ ~ N(0, 1^2^).

In the setting where there was instrument related error in the outcome, and pleiotropy (scenario 2A in Supplementary Figure 1), we simulated the observed, error prone, outcome as:

O’_i, p_ = O_p_ + I_2_ + ε_19_

where ε_19_ ~ N(0, 1^2^).

In the setting where there was instrument related error in the exposure, and pleiotropy (scenario 4A in Supplementary Figure 1), we used O_p_ as the outcome and E’_I_ as the observed exposure.

To simulate population structure (scenarios 2B and 4B in Supplementary Figure 1) in this setting we simulated the measurement error mechanism as:

N_I,ps_ = I_1_ + I_2,ps_ + ε_20_

where ε_20_ ~ N(0, 1^2^).

And the observed, error-prone, exposure as (scenario 4D in Supplementary Figure 1):

E’_I,ps_ = E + N_I,ps_ + ε_21_

where ε_21_ ~ N(0, 1^2^).

And the observed, error-prone outcome (scenario 2D in Supplementary Figure 1) as:

O’_I,ps_ = O_ps_ + N_I,ps_ + ε_22_

where ε_22_ ~ N(0, 1^2^).

To simulate the setting where we have only partially measured the mechanism through which the measurement error occurs (i.e. the measurement error is only partially mediated by the variable being adjusted for) (scenarios 2E and 4E in Supplementary Figure 1) we simulated the differential error in exposure and outcome as a function of both the mediator and a direct effect of the instrument:

E’_part, I_ = E + N_I_/2 + I_1_/2 + ε_23_

O’_part, I_ = O + N_I_/2 + I_1_/2 + ε_24_

where ε_23_ ~ N(0, 1^2^) and ε_24_ ~ N(0, 1^2^).

To explore the situations in which the mechanism is instead through a confounding effect (Scenarios in the F row of Supplementary Figure 1), rather than mediation, we simulated a mediator as

N_c_ = I_2_ + ε_25_

where ε_25_ ~ N(0, 1^2^).

In the setting where there is outcome related differential measurement error in the exposure (scenario 3F in Supplementary Figure 1) we simulated the true outcome as:

O_c1_ = E + C + N_c_ + ε_26_

where ε_26_ ~ N(0, 1^2^).

And, the observed, error prone, the exposure as

E’_e<-o,c_ = E + N_c_ + ε_27_

where ε_27_ ~ N(0, 1^2^).

In the setting where there is exposure related differential measurement error in the outcome (scenario 1F in Supplementary Figure 1), we simulated the true exposure as:

E_c2_ = I_1_ + N_c_ + C + ε_28_

where ε_28_ ~ N(0, 1^2^).

The true outcome as:

O_c2_ = E_c2_ + C + ε_29_

where ε_29_ ~ N(0, 1^2^).

And, the observed, error prone, outcome as

O’_e->o,c_ = O_c2_ + N_c_ + ε_30_

where ε_30_ ~ N(0, 1^2^).

In the setting where there is instrument related differential measurement error (scenarios 2F and 4F in Supplementary Figure 1), we simulated the instrument as:

I_1,c_ = N(0, 1^2^). + N_c_

When the error is in the outcome (scenario 2F in Supplementary Figure 1), we simulated the true exposure as:

E_c3_ = I_1,c_ + N_c_ + C + ε_31_

where ε_31_ ~ N(0, 1^2^).

The true outcome as:

O_c3_ = E_c3_ + C + ε_32_

where ε_32_ ~ N(0, 1^2^).

And, the observed, error prone, outcome as:

O’_I,c1_ = O_c3_ + N_c_ + ε_33_

where ε_33_ ~ N(0, 1^2^).

When the error is in the exposure (scenario 4F in Supplementary Figure 1), we simulated the true exposure as :

E_c4_ = I_1,c_ + C + ε_34_

where ε_34_ ~ N(0, 1^2^).

The true outcome as:

O_c4_ = E_c4_ + C + ε_35_

where ε_35_ ~ N(0, 1^2^).

And, the observed, error prone, exposure as:

E’_I,c1_ = E_c4_ + N_c_ + ε_36_

where ε_36_ ~ N(0, 1^2^).

*Estimands*: The estimand was again the average causal effect of the exposure on the outcome.

*Methods*: We used the two-stage least squares regression estimator from the ivreg package to generate the MR estimates. In the simulation of Figure 1, we calculated both a multivariable estimate and a univariable estimate, while the simulation of Supplementary Figure 1 calculated the MVMR estimate only. The univariate MR estimates were generated from a two-stage least square regression including the exposure (or, when appropriate, a miss measured proxy), its genetic risk score, and the outcome (or, when appropriate, a miss measured proxy). The model for the MVMR estimates additionally included the measurement error mechanism (or, when appropriate, a mismeasured proxy) and its genetic risk score. To aid interpretation, we additionally ran a univariate models for the situation in which the measurement error in exposure and outcome had a common cause, as well as our default setting in which the measurement error in outcome was caused by exposure (and vice versa).

*Performance measures*: Bias and 95% confidence interval coverage were again used as the performance measures.

**3. Simulations to explore the impact of phenotypic non-differential measurement error in the discovery sample on MR estimates.**

Aims: To explore the effect of phenotypic non-differential measurement error in the discovery sample of a GWAS (used to identify genetic instruments), we first simulated an MR study using a discovery GWAS with incrementally more measurement error.

*Data generating model*: The data generative model included 150 SNPs, each of which could take the values of 0, 1 or 2. The SNPs had a mean effect allele frequency of 0.5 (SD = 0.1), and 50 SNPs had an effect size of 0, 50 SNPs an effect size of 0.1, and 50 SNPs an effect size of 0.2. The exposure was then the product of each of the SNPs multiplied by their effect sizes, plus a confounder (modelled as a random normal variable with a mean of zero, SD of 1 and effect size of 1), and an error term with a mean of zero and a standard deviation of 4.7. The standard deviation of the error term was set so that the genotypes explained around 5% of the variance. The discovery GWAS had approximately 80% power for detecting a SNP effect size of 0.1 with 200,000 participants. The outcome was then a linear function of the exposure, confounder and an error term modelled as a random normal variable with a mean of zero and standard deviation of 1. Measurement error was then introduced to the exposure by adding an error term with a mean of zero and an incrementally larger standard deviation. Linear regression models were then run in a sample of 200,000 participants for each SNP to decide which SNPs were associated with the exposure at a 5x10^-8^ p-value threshold. This process was repeated 1000 times. The SNP-exposure and SNP-outcome associations were then derived from further linear regression models, each ran in independent samples with 200,000 participants.

More formally, we simulated all 150 SNPs each independently as the sum off two independent and identically distributed binomial variables with the following parameters:

P ~ N(0.5,0.1^2^)

SNP = B(1,P) + B(1,P)

The confounder was simulated as a Normal random variable:

C ~ N(0, 1^2^)

The exposure was then simulated as:

E = C + $\sum_{1}^{50} (0.1*SNP)$ + $\sum_{51}^{100} (0.2*SNP)$ + $\sum_{150}^{101} (0*SNP)$ + ε_1_

where ε is an error term such that ε_1_ ~ N(0, 4.7^2^). The error term was set so that the variance explained by all the SNPs is around 5%.

The measurement error in the exposure was then simulated as:

E* = E + ε_2_

where ε_2_ ~ N(0, i^2^) and i varied from 0.13 to 6.6.

The outcome was then simulated as:

O = E + C + ε_3_

where ε_3_ ~ N(0, 1^2^).

*Estimands*: precision and bias in the causal estimates, as well as the number of SNPs selected by the discovery sample.

*Methods:* We then recorded: A) the inverse-variance weighted (IVW) MR estimate when: 1) the same sample was used to estimate the SNP-exposure association for the MR analysis as was used to select the SNPs, 2) the same sample was used to estimate the SNP-exposure association for the MR analysis as was used to select the SNPs, but a winner’s curse correction (described below) was applied, and 3) the sample used to estimate the SNP-exposure association for the MR analysis was different from the one used to select the SNPs. B) the SNP-exposure regression coefficient for the SNPs used in the MR analysis estimated in 1) the same sample used to select the SNPs used in the MR analysis, 2) the same sample used to select the SNPs used in the MR analysis after the winner’s curse correction was applied, and 3) a separate sample from the sample used to select the SNPs used in the MR analysis.

*Performance measures*: Bias and size of the standard error (SE) in the SNP effect sizes and MR estimates, as well as the number of SNPs and precision of the IVW estimate.

**4. Power Analysis for the simulations**

No *a priori* power calculation was conducted to choose the number of repetitions run in each simulation.

**5. Inverse Quantile Transformation (FIQT) Winner’s curse correction**

The FIQT winner’s curse correction uses an analogy between multiple testing and Winner’s curse to create an easy-to-implement statistical correction to the SNP-effect estimates. Specifically, the FIQT runs a multiple test correction on the SNP p-value, and then they use this to get a corrected z-statistic for the SNP-exposure effect. Dividing this by the original z-statistic creates a deflation factor for the SNP-exposure effect estimate. Previous simulations have found that the FIQT reduced the mean squared error of GWAS summary statistics when compared to no correction.(5)

**6. Study design and data sources for the applied examples**

UK Biobank (UKB)

Setting

The UK Biobank (UKB) is a cohort study of people living in England, Scotland and Wales. UKB participants were recruited members of the public aged 38-73 who lived within 22 miles of an assessment centre between 2006 and 2010. Overall, 503,325 individuals participated in the study, with a total response rate of approximately 6%. The full protocol is available online.(6) The study design, participants and quality control (QC) methods have been described in full elsewhere.(7)

GWAS methods

All UKB GWASs were conducted using the MRC-IEU UKB GWAS pipeline.(8) A full description of the pipeline methods, including Quality Control filtering and imputation, can be found in the original citation. In brief, multiallelic Single Nucleotide Polymorphisms (SNPs) or those with minor allele frequency ≤0.01 were removed. The remaining SNPs were imputed using the UK10K haplotype and Haplotype Reference Consortium reference panels. Additionally, sex-mismatched or sex-chromosome aneuploidy individuals were excluded. Previously derived association statistics were then created using a linear mixed model implemented in BOLT-LMM,(9) adjusting for sex and SNP-chip. The weight GWAS run specifically for this study used BOLT-LLM and adjusted for first ten genetic principal components, age, sex, and chip.(8)

Participants

One sample MR used people who had provided genetic data and had not withdrawn consent (N = 462885). All UKB samples included both men and women and participants were predominantly of European ancestry.

Phenotypic measures

Glucose (UKB ID: 30740, OpenGWAS ID: met-d-Glucose, GWAS N: 114,867) was measured by hexokinase analysis on a Beckman Coulter AU5800 at the initial assessment visit, with units of mmol/L.

Time since last eating (UKB ID: 74, OpenGWAS ID: ukb-b-16156, GWAS N: 462,992) was defined as the time interval between consumption of food or drink and blood sample(s) being taken.

Food weight (UKB ID: 100001, OpenGWAS ID: ukb-b-13330, GWAS N: 64,979) was defined as the estimated weight of all food and beverages consumed the previous day (excluding supplements) and was asked via a questionnaire during an online follow-up.

BMI (UKB ID: 21001, OpenGWAS ID: ukb-b-19953) was derived from the height and weight measurements taken at the initial assessment visit. The variable was standardised, with one standard deviation representing 4.8 Kg/m^2.

Weight (UKB ID: 21002, OpenGWAS ID: ukb-b-11842, GWAS N: 461,632) was measured at the initial assessment visit, with units of Kg.

Hair colour [used as a negative control outcome for residual population structure in the BMI applied example] (UKB ID: 1747) was assessed through the question:  "What best describes your natural hair colour? (If your hair colour is grey, the colour before you went grey)", during baseline assessment. 6 GWASs were used: black hair (OpenGWAS ID: ukb-d-1747_5, N case: 15,809, N control: 344,461), dark brown hair (OpenGWAS ID: ukb-d-1747_4, N case: 134,627, N control: 225,643), light brown hair (OpenGWAS ID: ukb-d-1747_3, N case: 147,560, N control: 212,710), blonde (OpenGWAS ID: ukb-d-1747_1, N case: 41,178, N control: 319,092), red (OpenGWAS ID: ukb-d-1747_2, N case: 16,615, N control: 343,655), and other (OpenGWAS ID: ukb-d-1747_6, N case: 4,481, N control: 355,789).

Age and sex (UKB ID: 21022 and 31 respectively) were asked through questionnaires at the baseline assessment.

Ethics

UKB received ethical approval from the North West Multi-Centre Research Ethics Committee (REC reference 11/NW/0382). All participants provided written informed consent to participate in the study. Data from the UKB are fully anonymised.

MAGIC 2013 (10)

Participants

The study was a meta-analysis that had 133,010 participants, of European ancestry including both males and females. Studies were asked, if applicable, to adjust for age, study site, and geographical covariates. Details of the participating studies can be found in the original publication.

GWAS methods

Participating studies genotyped participants using an Illumina array, applied QC metrics and imputed SNPs using the HapMap. More details are provided in the original publication.

Phenotypic measures

Fasting glucose (OpenGWAS ID: ieu-b-113) was measured in serum with a mmol/l unit. Fast time varied for study to study, but was generally either at least 8 hours or overnight.

GIANT 2015 (11)

Participants

The study was a meta-analysis of 339,224 participants, drawn from a mixed ethnicity population, including both males and females. Of the 339,224 participants, 322,154 were of European descent. Details of the participating studies can be found in the original publication.

GWAS methods

Participating studies genotyped participants using an Affymetrix or Illumina array, applied QC metrics and imputed SNPs using the HapMap. More details are provided in the original publication.

Phenotypic measures

BMI (OpenGWAS ID: ieu-a-2) was defined as the kg per m^2, but was standardised in the meta-analysis used in this study.

FinnGen (12,13)

Setting

FinnGen is a research project which includes nine Finnish biobanks, universities, and university hospitals, and 13 international pharmaceutical industry partners. The project links genomic data in up to 224,737 individuals (release 5) with nationwide health register data collected from every resident in Finland since 1969..

Participants

The sample included 111,108 cases and 107,684 controls, from a Finnish population, including both males and females.

GWAS methods

Genotyping used Illumina and Affymetrix arrays. GWAS models adjusted for age, sex, the first 10 genetic principal components, and genotyping batch. Further details about the GWAS methods, including QC procedure, can be found on the documentation on their website.(14)

Phenotypic measures

Cardiovascular Disease (CVD) (OpenGWAS ID: finn-b-I9_CVD) was derived from medical data linkage. More details can be found on the FinnGen website.(12) Because the GWAS used logistic regression, beta weights from this GWAS represent the log odds of CVD risk.

GIANT 2013 (15)

Participants

The study was a meta-analysis of 73,137 female and 60,586 male participants, drawn from a European population. Details of the participating studies can be found in the original publication. There was no sample overlap with FinnGen. In OpenGWAS, the data from this study was provided as two sex-specific GWASs (i.e. one male only GWAS, and one female only GWAS). We combined these two GWASs by averaging SNP effect size of each sex’s GWASs. This estimate should be equivalent to the sex-adjusted effect estimate.

GWAS methods

Participating studies genotyped participants using an Affymetrix or Illumina array. Details about QC and imputation are provided in the original publication.

Phenotypic measures

Weight (OpenGWAS ID: ieu-a-107 for females, and ieu-a-106 for males) was defined in the SD scale, and was transformed into the Kg scale by multiplying the effect estimates by the standard deviation of each scale (i.e. 13.57 and 13.36 respectively for females and males).

**7. Assumptions of MR**

All Mendelian randomisation studies make three core (instrumental variables) assumptions (16):

1) Relevance: that the variant is a reliable predictor of the exposure. The rule of thumb for this assumption is that the F statistic for the variant-exposure association should be ten or greater.
2) Independence: that there are no variant-outcome confounders. The most common threat to this assumption in MR is confounding through population structure, which is less likely when the outcome GWASs used a linear mixed model or adjusted for genetic principal components.
3) Exclusion restriction: that the variants cause the outcome only through the exposure. In genetics, the most common violation of this assumption is pleiotropy. Because pleiotropic effects should be different for every SNP, pleiotropy should result in heterogeneity in the MR estimates which allows for a weakening of this assumption in a two-sample setting.

Two sample MR additionally assumes that the two samples are taken from the same population but do not have any overlap. Sample overlap was estimated by manually searching the supplementary materials of the included consortia for which samples and the number of participants included. The maximum potential overlap was calculated as the smaller number of participants included in either consortium for every overlapping study.

**8. Statistical Methods for the applied examples**

Selection of genetic variants

The anthropometric (BMI and weight) instruments were selected based on a threshold of p < 5x10^-8, while the consumption (time since last meal and food weight) instruments were selected based on a threshold of p < 5x10-6. BMI instruments were selected from the GIANT 2015 sample. SNPs were then clumped based on an r^2 of 0.001 within a 10000kb window.

MR assumes that the causal path goes from variant to exposure to outcome, not variant to outcome to exposure. In two-sample MR analyses where both the exposure and outcome were continuous (i.e. the fasting/non-fasting glucose example) we used Steiger filtering to remove SNPs which have a stronger correlation with the outcome than the exposure (implying that the exposure is more distal than the outcome). We used the default values in the harmonise_data() function in TwoSampleMR package version 0.5.6 to harmonise data, and to keep palindromes when the minor allele frequency can be used to infer the positive strand.

MR estimators

In the one sample MR analysis, the MR-estimator was the beta parameter from a two-stage least squared regression. This parameter is equivalent to the beta parameter from a regression using the outcome as the dependent variable and the values of the exposure which are predicted by the instrument as an independent variable.

The primary two-sample estimator was the Wald ratio, which is the variant-outcome association divided by the variant-exposure association. The primary estimator method of aggregating this ratio across SNPs was Inverse Variance weighting (IVW). This will produce valid weights when all MR assumptions are valid. In addition, for the applied example looking at the effect of BMI on fasting blood glucose, we additionally implemented:

1) MR-Egger, a method which will provide valid estimates in the presence of pleiotropy assuming that the strength of the variant-exposure association is independent of the pleiotropic association between the SNP and the outcome. A common violation of this assumption is when the pleiotropic effect occurs through a confounder of the exposure-outcome association. MR-Egger also assumes that there is no measurement error in the exposure (NOME) GWAS from which the weights were derived as an alternative to the instrumental variables assumption that there are no weak instruments. An I^2 for the variant-exposure association is greater than 90% is viewed as the NOME equivalent of an F statistic of 10.

2) Weighted median method, which gives a consistent causal estimate assuming that at least half of the weights come from valid instruments.

3) The weighted mode method, which assumes that the plurality/mode of the Wald ratio effects is a valid estimate of the true effect, often dubbed the Zero Modal Pleiotropy Assumption (ZEMPA). The weighted median and weighted mode methods weight the Wald ratios by the inverse of the variance in the variant-outcome association and therefore also make the NOME assumption.

Missing data

We did not impute any missing data in the individual-level data analyses. However, missing SNPs (including palindromes) were imputed in the two sample MR analyses when they had an LD r^2 of 0.8 and a MAF > 0.3, based on the 1000 Genomes European reference panel.

**9. Creation of Genetic Risk Scores (GRS) and one sample estimation in applied examples**

**Genetic Risk Scores (GRS) and Correlations (rG)**

We additionally explored the extent to which the presence of this differential measurement error may bias the estimated genetic correlation. To do this we created genetic risk scores (GRSs) for the three GWAS above using the MRBase-GRS package.(17) The GRS included only SNPs associated with each trait at p < 5x10^-8^, clumped at r^2^ of 0.001 and within a genetic distance of 10,000 kb. The genetic correlations were then calculated using individual-level data in the UKB, after removing participants without genetic data or who had withdrawn consent, as the partial correlation of the BMI GRS with the two glucose GRSs after adjusting for the first 40 principal components (PCs) of genetic ancestry, age, sex, and genotyping chip.

**One Sample MR modelling**

In this analysis, we created a weighted genetic risk score (GRS) for BMI using the GIANT GWAS used above. We then performed a one-sample MR analysis in the UKB, using the GRS as an instrument for BMI (the primary exposure), and non-fasting blood glucose as the outcome. Because drinking water may not necessarily influence blood glucose levels in the same way as eating food, we used MVMR to adjust for both self-reported food or drink consumption and another question, on food weight eaten the previous day, to reduce the possibility that we only partially adjust for food consumption. The GWAS summary statistics used to create the instruments for these secondary exposures were created in UKB and extracted from the MRC-IEU OpenGWAS platform.(18) Because neither GWAS had any genome-wide significant SNPs we instead used a 5x10^-6^ p-value for instrument construction. By combining the SNPs into a GRS in a one-sample setting, we could maintain instrument strength despite using non-genome-wide significant SNPs.

After excluding participants without genetic data or who had withdrawn consent, we ran a two-stage least square regression model in the UKB with the GAINT 2015 GRS for BMI and the two food consumption variables’ GRSs as the instruments, BMI and the two food consumption variables as exposures, non-fasting blood glucose levels as the outcome, and age, sex, genotyping chip, and the first 40 genetic PCs as covariates. We additionally calculated the total MR estimate in a one-sample setting by running the same analysis without the food consumption variables and their GRSs.

**10. Assessment of assumptions in applied examples**

The F statistics in the two sample MR analyses were calculated by squaring the beta value for the variant-exposure association and dividing it by the square of the standard error of that association. In the one sample setting, they were taken from the output of ‘ivreg’. In a one sample setting we tried to remove any effect of population structure by adjusting for the first 40 genetic principal components, while in the two sample setting we check that the GWASs had adjusted for genetic principal components or used a linear mixed model. To check that the samples are drawn from the same population we check that they use mostly participants from the same ancestry and include the same sexes. For the analysis of the effect of BMI on blood glucose we additionally used heterogeneity statistics to explore the existence of pleiotropy in two sample MR analyse.

**11. Sensitivity and additional analyses in applied examples**

We used hair colour as a negative control outcome for the instruments used in the applied example of the effect of BMI on blood glucose. Because hair colour should not associate with glucose, but does vary based on ancestry, we used it to test for residual confounding from population structure.(19) Because a pleiotropic effect would still be evidence of residual confounding in the instruments, we only use the IVW estimator for this analysis.

**12. Software and preregistration**

MR analyses in this paper were run using the TwoSampleMR R package and the ivreg R package.(3,4) The genetic correlations were calculated using the ppcor package.(20) The GRSs were created using the MR-Base GRS package.(17) Graphs were drawn using ggplot2 and DAGitty.(21,22) Most GWAS data was extracted from the IEU OpenGWAS platform.(18) The conditional f statistic was calculated using the OneSampleMR r package.(23) This study was written in accordance with STROBE-MR and the ADEMP guidelines.(1,24) The study was not pre-registered

**References**

1.	Morris TP, White IR, Crowther MJ. Using simulation studies to evaluate statistical methods. Statistics in Medicine. 2019;38(11):2074–102.

2.	Hernán MA, Robins JM. Causal Inference: What If. :311.

3.	Hemani G, Zheng J, Elsworth B, Wade KH, Haberland V, Baird D, et al. The MR-Base platform supports systematic causal inference across the human phenome. eLife. 2018 May 30;7:e34408.

4.	Fox J, Kleiber C, Zeileis A, Kuschnig N. ivreg: Instrumental-Variables Regression by “2SLS”, “2SM”, or “2SMM”, with Diagnostics [Internet]. 2021 [cited 2022 Apr 6]. Available from: https://CRAN.R-project.org/package=ivreg

5.	Bigdeli TB, Lee D, Riley BP, Vladimirov V, Fanous AH, Kendler KS, et al. FIQT: a simple, powerful method to accurately estimate effect sizes in genome scans [Internet]. bioRxiv; 2015 [cited 2022 Mar 12]. p. 019299. Available from: https://www.biorxiv.org/content/10.1101/019299v1

6.	Collins R. What makes UK Biobank special? Lancet. 2012 Mar 31;379(9822):1173–4.

7.	Ruth Mitchell E. MRC IEU UK Biobank GWAS pipeline version 2 [Internet]. data.bris. 2019 [cited 2022 Mar 12]. Available from: https://data.bris.ac.uk/data/dataset/pnoat8cxo0u52p6ynfaekeigi

8.	Loh PR, Tucker G, Bulik-Sullivan BK, Vilhjálmsson BJ, Finucane HK, Salem RM, et al. Efficient Bayesian mixed-model analysis increases association power in large cohorts. Nat Genet. 2015 Mar;47(3):284–90.

9.	Loh PR, Kichaev G, Gazal S, Schoech AP, Price AL. Mixed-model association for biobank-scale datasets. Nat Genet. 2018 Jul;50(7):906–8.

10.	Scott RA, Lagou V, Welch RP, Wheeler E, Montasser ME, Luan J, et al. Large-scale association analyses identify new loci influencing glycemic traits and provide insight into the underlying biological pathways. Nat Genet. 2012 Sep;44(9):991–1005.

11.	Locke AE, Kahali B, Berndt SI, Justice AE, Pers TH, Day FR, et al. Genetic studies of body mass index yield new insights for obesity biology. Nature. 2015 Feb 12;518(7538):197–206.

12.	FinnGen. FinnGen Documentation of R5 release. 2021; Available from: https://finngen.gitbook.io/documentation/v/r5/

13.	Kurki MI, Karjalainen J, Palta P, Sipilä TP, Kristiansson K, Donner K, et al. FinnGen: Unique genetic insights from combining isolated population and national health register data [Internet]. medRxiv; 2022 [cited 2022 Apr 14]. p. 2022.03.03.22271360. Available from: https://www.medrxiv.org/content/10.1101/2022.03.03.22271360v1

14.	Risteys FinnGen R6 - I9_CVD [Internet]. [cited 2022 Apr 6]. Available from: https://r6.risteys.finngen.fi/phenocode/I9_CVD

15.	Randall JC, Winkler TW, Kutalik Z, Berndt SI, Jackson AU, Monda KL, et al. Sex-stratified Genome-wide Association Studies Including 270,000 Individuals Show Sexual Dimorphism in Genetic Loci for Anthropometric Traits. PLOS Genetics. 2013 Jun 6;9(6):e1003500.

16.	Sanderson E, Glymour MM, Holmes MV, Kang H, Morrison J, Munafò MR, et al. Mendelian randomization. Nat Rev Methods Primers. 2022 Feb 10;2(1):1–21.

17.	sean-harrison-bristol. UK_Biobank_PRS [Internet]. 2021 [cited 2022 Apr 6]. Available from: https://github.com/sean-harrison-bristol/UK_Biobank_PRS

18.	Elsworth B, Lyon M, Alexander T, Liu Y, Matthews P, Hallett J, et al. The MRC IEU OpenGWAS data infrastructure [Internet]. bioRxiv; 2020 [cited 2022 Mar 30]. p. 2020.08.10.244293. Available from: https://www.biorxiv.org/content/10.1101/2020.08.10.244293v1

19.	Sanderson E, Richardson TG, Hemani G, Davey Smith G. The use of negative control outcomes in Mendelian randomization to detect potential population stratification. International Journal of Epidemiology. 2021 Aug 1;50(4):1350–61.

20.	Kim S. ppcor: An R Package for a Fast Calculation to Semi-partial Correlation Coefficients. Commun Stat Appl Methods. 2015 Nov;22(6):665–74.

21.	Wickham H. ggplot2. WIREs Computational Statistics. 2011;3(2):180–5.

22.	Textor J, van der Zander B, Gilthorpe MS, Liśkiewicz M, Ellison GT. Robust causal inference using directed acyclic graphs: the R package ‘dagitty.’ International Journal of Epidemiology. 2016 Dec 1;45(6):1887–94.

23.	Palmer T. R and Stata packages for one-sample Mendelian randomization analyses: OneSampleMR and ivonesamplemr. :20.

24.	Skrivankova VW, Richmond RC, Woolf BAR, Davies NM, Swanson SA, VanderWeele TJ, et al. Strengthening the reporting of observational studies in epidemiology using mendelian randomisation (STROBE-MR): explanation and elaboration. BMJ. 2021 Oct 26;375:n2233.
