## Supplementary Results for "Re-evaluating the robustness of Mendelian randomisation to measurement error"

**1. Simulations exploring the effect of differential measurement error in the exposure or outcome on MR estimates.**

Differential measurement error biased MR estimates and reduced coverage when error in the outcome was associated with the exposure or instrument (Supplementary Table 3). Similar findings were found when the measurement error in the exposure was associated with the outcome or the instrument and there was a true causal effect. However, when differential error in the exposure measure was associated with either the genetic variant or the outcome but the null hypothesis was true, the MR estimates were not biased and there was no reduction of coverage. In our simulations, after adjusting for all confounders, the average bias in the observational and MR estimates were similar. The two exceptions to this were when the exposure was measured with differential error under the null. In this setting, unlike MR, the observational estimates were biased.

**2. Change in the (genetic) correlation under differential measurement error.**

Simulation: Similar results were found for the correlations as were seen for the MR estimates. For example, in the same setting, the phenotypic correlation was biased by 0.146 (Monte Carlo 95%CI 0.146 to 0.146) while the genetic correlation was biased by 0.187 (Monte Carlo 95%CI 0.187 to 0.187). However, unlike MR, the genetic correlation was not biased by any instrument related measurement error in the exposure (Supplementary Table 6).

Applied example of the effect of BMI on fasting/non-fasting blood glucose: The genetic correlation between BMI and blood glucose was halved from -0.024 (SE = 10^-7, p < 0.001, for non-fasting) to -0.012 (SE = 10^-7, p < 0.001, for fasting blood glucose).

**3. Two-sample Mendelian randomisation (MR) applied example outputs.**

The variant-exposure and variant-outcome associations for body mass index (BMI) on non-fasting glucose can be found in Supplementary Table 7. The variant-outcome and variant-exposure associations for the BMI single-nucleotide polymorphisms (SNPs) on fasting glucose can be found in Supplementary Table 8. The variant-exposure and variant-outcome associations for the effect of weight on cardiovascular disease (CVD) risk, using a discovery genome-wide association study (GWAS) with no measurement error, can be found in Supplementary Table 9. The final MR output for the effect of BMI on fasting/non-fasting blood glucose can be found in Supplementary Table 10.

**4. Additional sensitivity analysis for the applied examples.**

Without any correction for multiple tests, BMI associated with blonde and red hair; fasting with black hair (Supplementary Table 11). This implies that there may be some residual population structure (i.e. that ancestry or a variable associated with it is a confounder of the instrument-outcome association) in the instruments.

The F and I^2 statistics for the BMI SNP associations can be found in Supplementary Tables 7 and 8. Overall, fasting glucose had a mean F-statistic of 67.72 and non-fasting had a mean of 65.76. I^2 statistics for both the variant-exposure associations were >0.9. In the one sample setting, the F statistic for the univariate analysis is 7308.58, and the conditional F statistic in the 1-sample- multivariable MR was 166.66 for BMI, 12.98 for time since last consumption, and 49.55 for estimated weight of food consumed on the previous day.

**5. The impact of phenotypic non-differential measurement error in the discovery sample on MR estimates.**

We found that increasing measurement error in the GWAS discovery sample reduced the number of SNPs selected in the discovery sample in both our simulated data and the applied example in the UKB of the effect of weight on CVD (Supplementary Figure 2), which increased the standard error of the MR IVW estimates (Supplementary Figure 3). For example, in the real data setting, we observed a fivefold increase in the SE for the effect of weight on CVD risk when the SNP-exposure associations were estimated in a separate sample from that used to select the SNP, from a mean of 0.042 to 0.186, as percentage increase in the exposure’s standard deviation due to measurement error increased from zero to 227%.

For the simulated data, increasing measurement error in the GWAS discovery sample introduced Winner’s curse and biased SNP-effect estimates away from the null (Supplementary Figure 4). For example, when measurement error had caused a 173% increase in the exposure’s standard deviation there was a mean bias of 0.005 (Monte Carlo SE = 0.00010). This was attenuated by using a non-overlapping replication sample as a source of SNP-exposure associations in the MR analysis (mean bias = 0.002, Monte Carlo SE = 0.00007), and partially attenuated by the FIQT Winners Curse correction (mean bias = 0.002, Monte Carlo SE = 0.00009).

As measurement error increased, the F statistic for the SNP-exposure association in the replication sample also increased (Supplementary Figure 5). Because no false positive SNPs were included as measurement error increased (Supplementary Figure 6), this would have occurred because the increasing measurement error resulted in lower power to detect less strongly associated SNPs. Hence, weak instrument bias was reduced as measurement error increased inversely proportional to the F-statistic in the IVW estimates when using a three sample MR design (Figure 4). Thus, given an F of 107 in the no-error GWAS in the simulated data, we should expect to see the ~0.9% deflation observed. On the other hand, when the estimates were derived from the discovery GWAS they were additionally biased by Winner’s curse (e.g., when there was a 173% increase in SD, mean bias = -0.028 Monte Carlo SE = 0.001). This was attenuated by the FIQT Winner’s curse correction (mean bias = -0.001 Monte Carlo SE = 0.001).

There was much less evidence of inflation of the SNP-exposure estimates in the UKB for any design (Supplementary Figure 4), and hence much less difference in the IVW estimates (Figure 4) for our applied example. The F statistics derived from the setting in which the SNP-exposure associations were derived from the discovery sample (with or without application of the FIQT Winners curse correction, and in both simulated and UKB data) were deflated when compared to the F statistic derived from the replication sample (Supplementary Figure 5).

**6. Additional comments on the MVMR simulation**

Although the mean bias induced by weak instrument bias was not huge, the inflated standard error of the bias is compatible with these violations introducing weak instrument bias. We additionally found that, when the measurement error mechanism was a confounder rather than a mediator, bias was not induced when there was error in the exposure (or outcome) due to the outcome (or exposure). Bias was, however, introduced if the error in the exposure (or outcome) was due to a confounder of the instrument [Supplementary Table 12]. The inflated standard error for bias when trying to remove differential error in the exposure due to the outcome when the mechanism is though a confounder is again an indicator that weak instrument bias has been introduced.

**7. Additional comments on the MVMR applied example**

A putative explanation for the different point estimates in the 1SMR and 2SMR analyses (i.e. the effects of 0.179 and 0.103 mmol/L respectively) are the different effects of weak-instrument bias and Winner’s curse in the two settings. Although the instruments in the 2SMR setting both appear strong (F-statistics of 66 and 68 respectively), weak instrument bias in a two-sample setting should be towards the null. On the other hand, in a one-sample MVMR analysis, weak instrument bias can bias in any direction, and the instruments in this setting were less strong (*F* = 167 for BMI, 13 for time since last consumption, and 50 for food weight).

| Simulated Data | UKB Data |
| --- | --- |
| 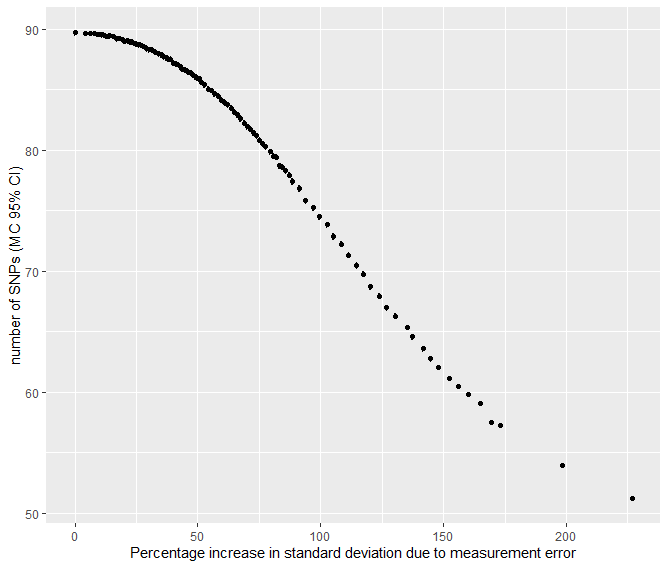 | 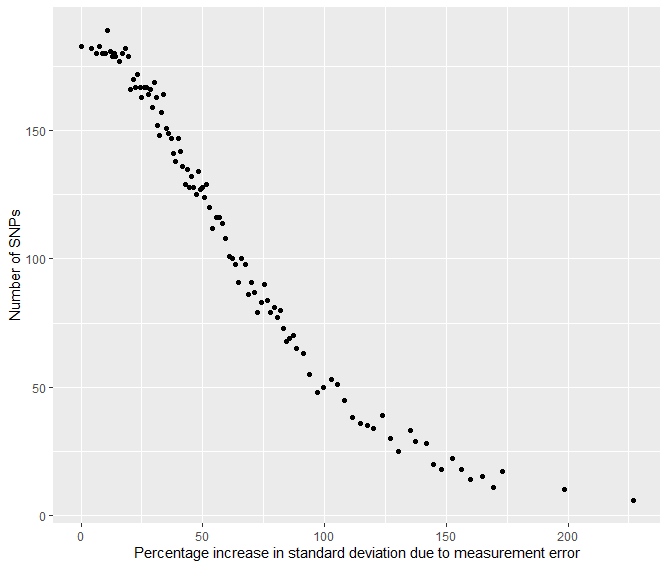 |

Supplementary Figure 2: The effect of measurement error on the number of SNPs selected in the discovery sample when using a p < 5x10^-8^ p-value threshold. SNP: single-nucleotide polymorphism; MC: Monte Carlo; CI: confidence interval.

|  | Simulated Data | UKB Data |
| --- | --- | --- |
| Discovery sample only | 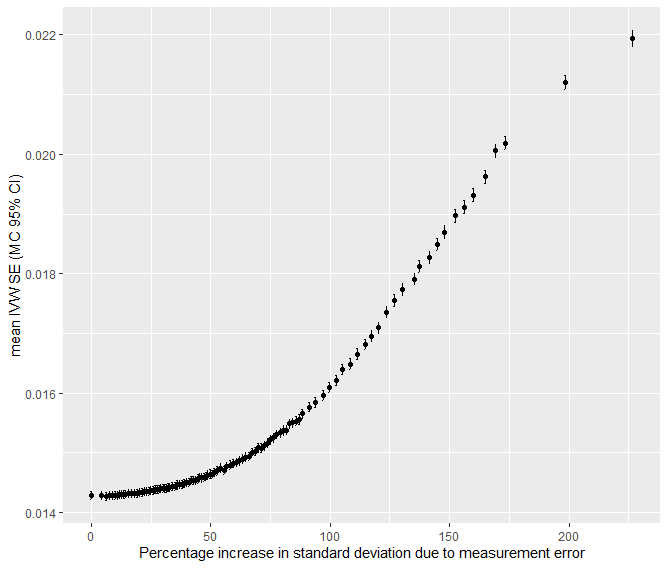 | 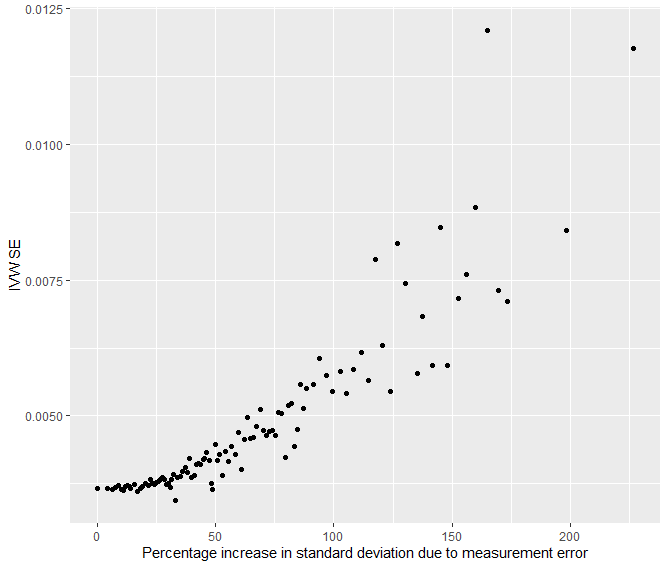 |
| Discovery sample after FIQT WCC | 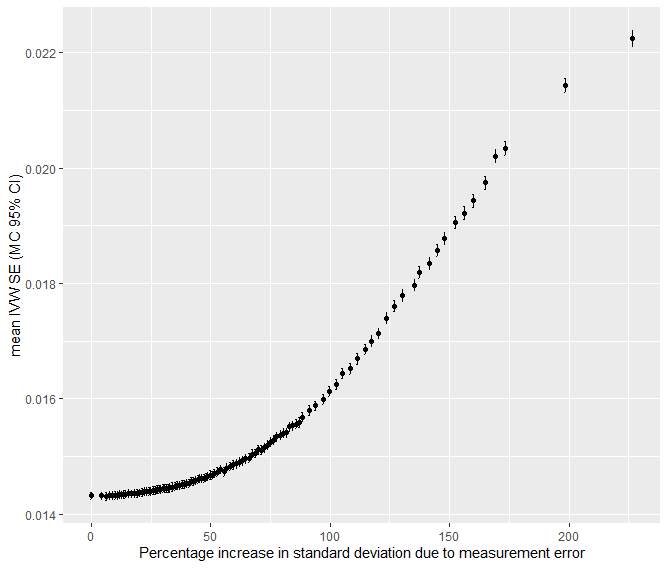 | 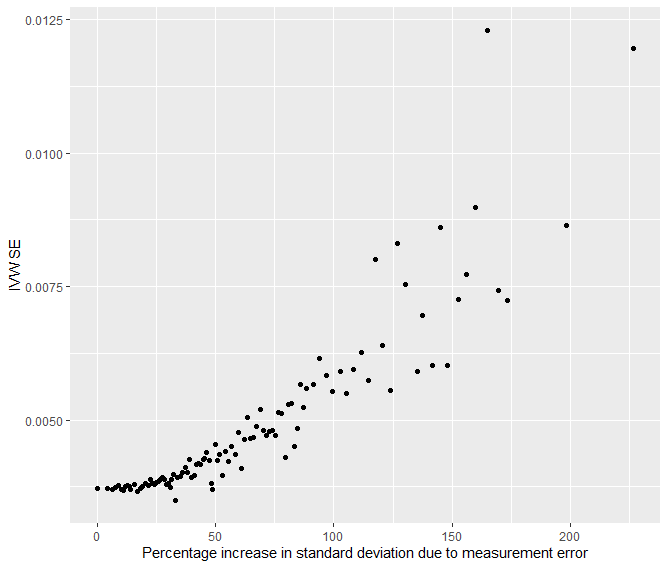 |
| Replication sample | 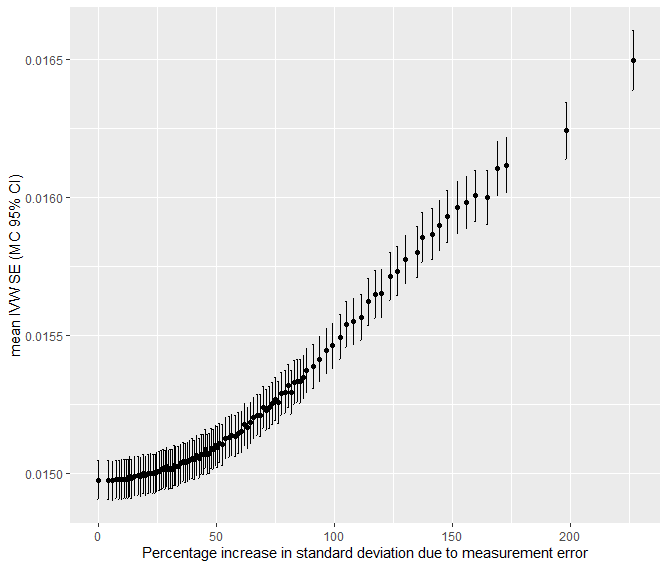 | 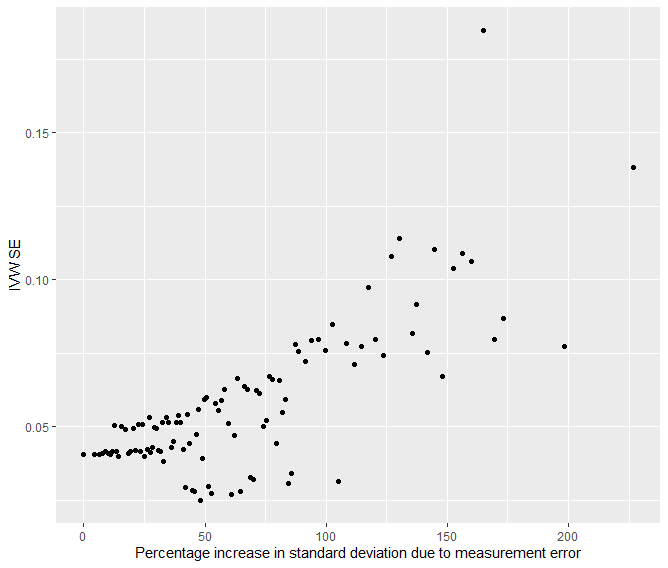 |

Supplementary Figure 3: The impact of non-differential measurement error in the discovery sample on standard error (SE) of the inverse-variance weighted (IVW) Mendelian randomisation estimates. CI: confidence interval; FIQT WCC: False discovery rate Inverse Quantile Transformation Winners Curse Correction.

|  | Simulated Data | UKB Data |
| --- | --- | --- |
| Discovery sample only | 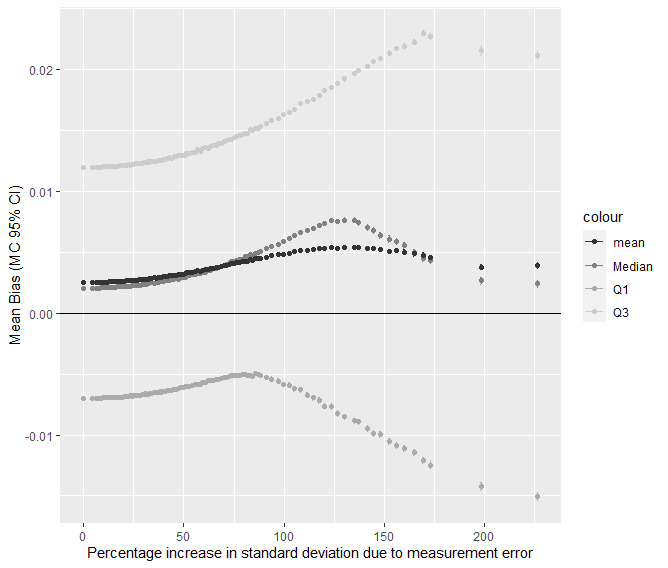 | 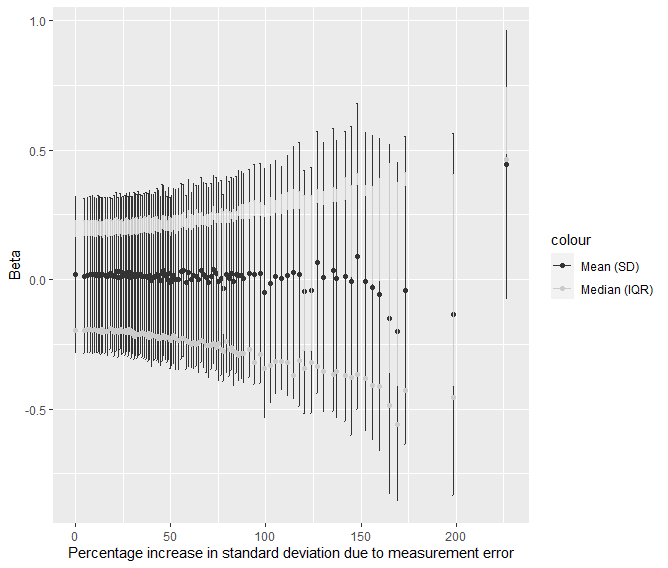 |
| Discovery sample after FIQT WCC | 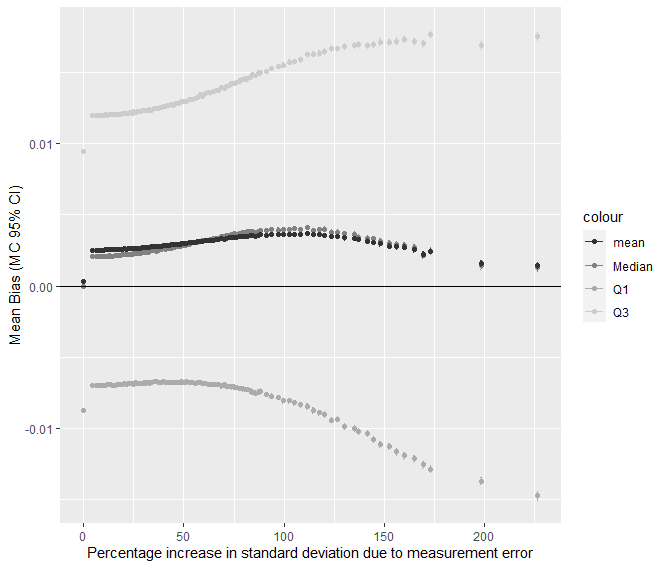 | 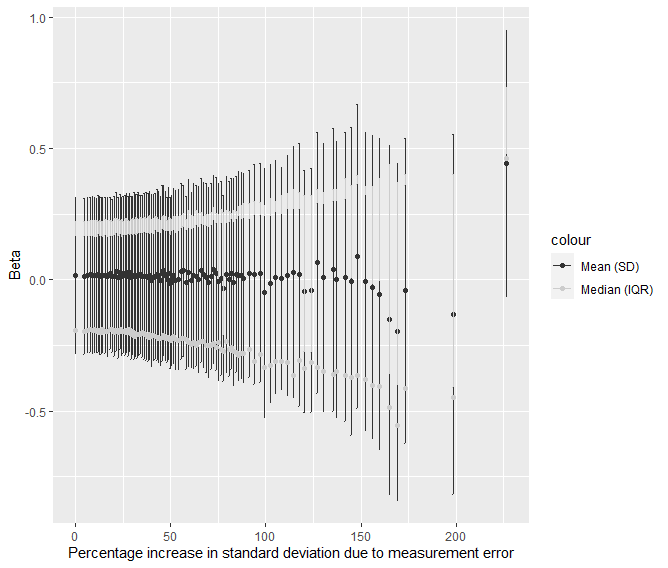 |
| Replication sample | 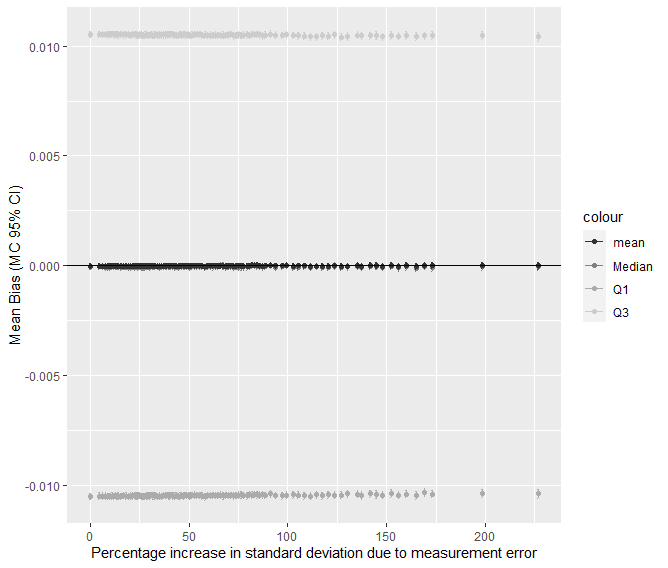 | 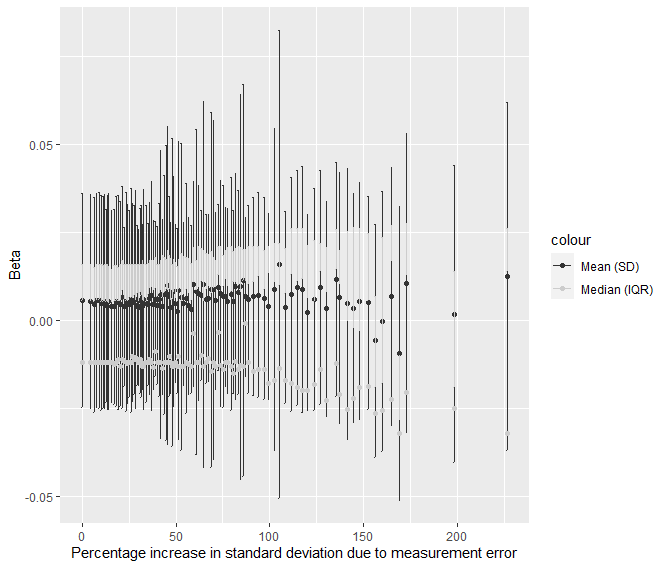 |

Supplementary Figure 4: Distribution of sizes of the SNP-exposure association. Left hand panel shows synthetic data with the y-axis plotting bias, while the right hand panel shows the raw effect sizes from the UKB. The first row (“Discovery sample only”) is the distribution of the SNP effect sizes selected as instruments when using the dataset for instrument selection and effect size estimation. The second row (“Discovery sample after FIQT WCC”) is the distribution of the SNP effect sizes selected as instruments when using the dataset for instrument selection and effect size estimation after apply the False discovery rate Inverse Quantile Transformation Winners Curse Correction. The bottom row (“Replication sample”) is the distribution of the SNP effect sizes selected as instruments when using different datasets for instrument selection and effect size estimation. GWAS: genome-wide association study; Q: quartile; MC: Monte Carlo; CI: confidence interval; SD: standard deviation; IQR: interquartile range.

|  | Simulated Data | UKB Data |
| --- | --- | --- |
| Discovery sample only | 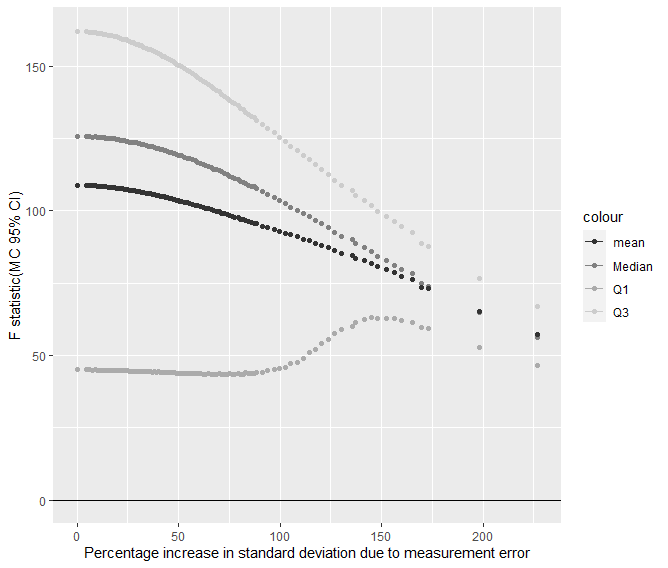 | 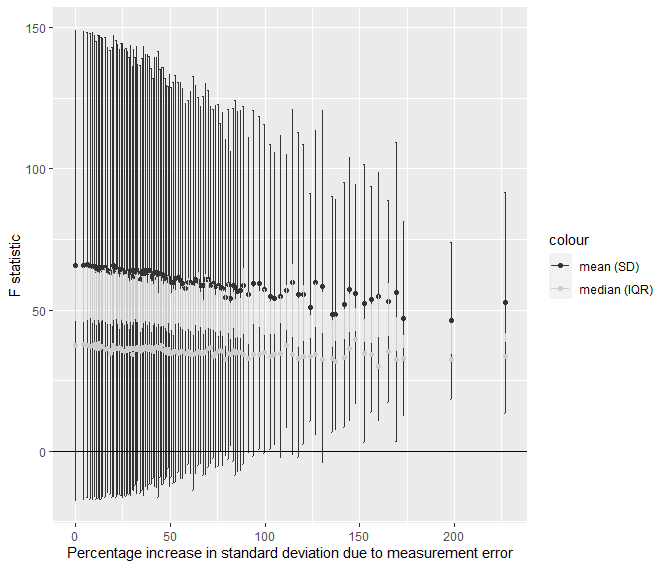 |
| Discovery sample after FIQT WCC | 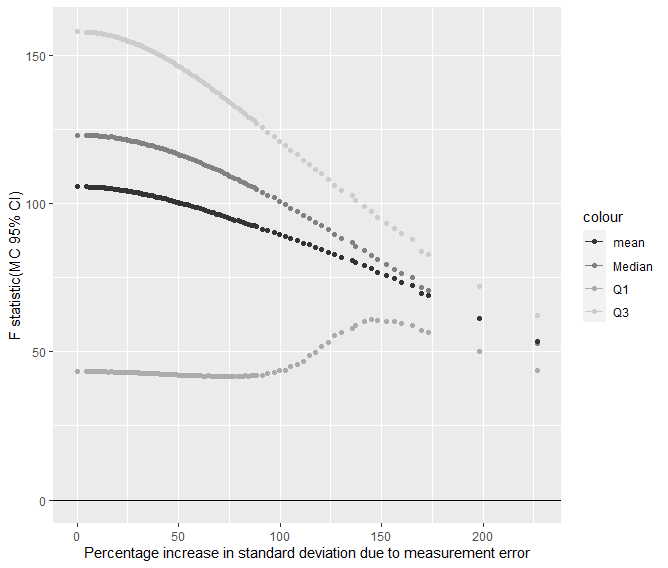 | 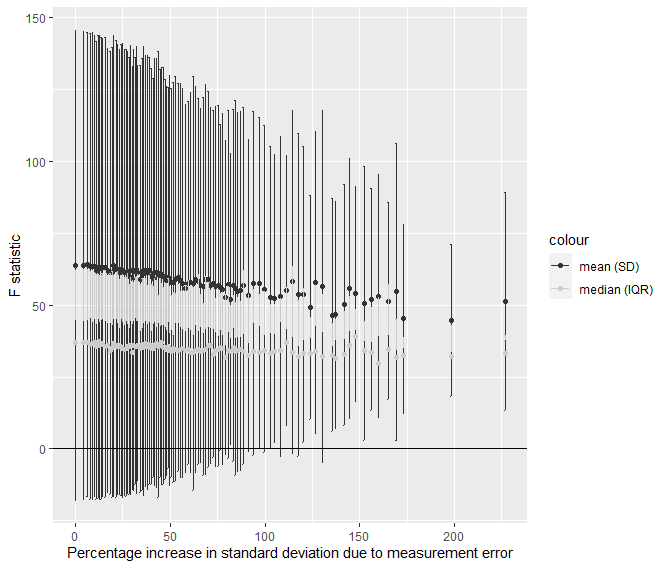 |
| Replication sample | 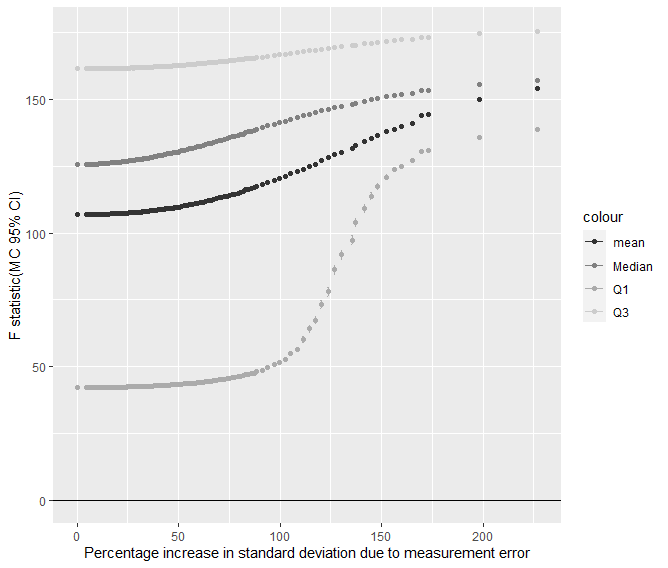 | 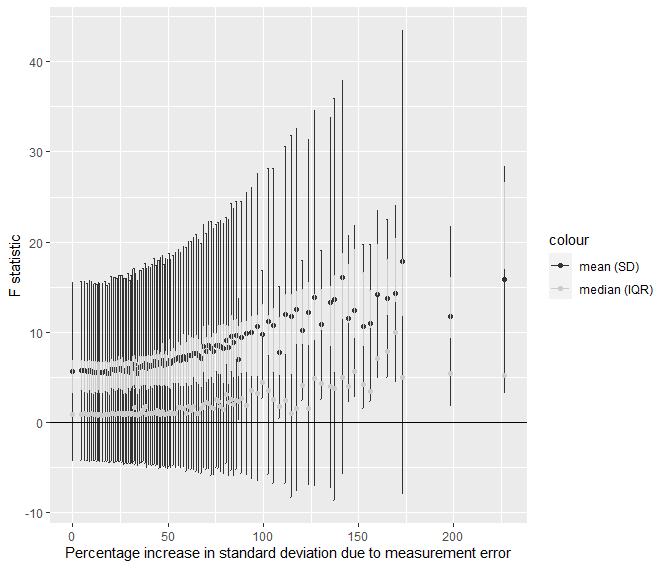 |

Supplementary Figure 5: Change in the distribution of the SNP-Exposure F-Statistic in the replication sample. MC: Monte Carlo; CI: confidence interval; SD: standard deviation; IQR: interquartile range; FIQT WCC: False discovery rate Inverse Quantile Transformation Winners Curse Correction.

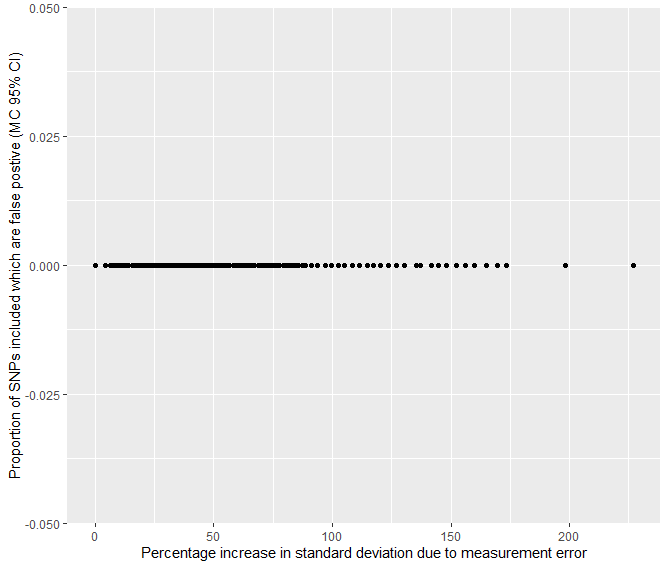

Supplementary Figure 6: Change in the number of false positive SNPs selected by the discovery sample in the simulated data as non-differential measurement error increased.
